# Tracking the temporal variation of COVID-19 surges through wastewater-based epidemiology during the peak of the pandemic: a six-month long study in Charlotte, North Carolina

**DOI:** 10.1101/2021.09.23.21258047

**Authors:** Visva Bharati Barua, Md Ariful Islam Juel, A. Denene Blackwood, Thomas Clerkin, Mark Ciesielski, Adeola Julian Sorinolu, David A. Holcomb, Isaiah Young, Gina Kimble, Shannon Sypolt, Lawrence S. Engel, Rachel T. Noble, Mariya Munir

**Author notes:** <u>Corresponding author:</u> Dr. Mariya Munir. These authors contributed equally (Visva Bharati Barua, Md Ariful Islam Juel).

## Abstract

The global spread of SARS-CoV-2 has continued to be a serious concern after WHO declared the virus the causative agent of the coronavirus disease 2019 (COVID-19) a global pandemic. Monitoring of wastewater is a useful tool for assessing community prevalence given that fecal shedding of SARS-CoV-2 occurs in high concentrations by infected individuals, regardless of whether they are asymptomatic or symptomatic. Using tools that are part of the wastewater-based epidemiology (WBE) approach, combined with molecular analyses, wastewater monitoring becomes a key piece of information used to assess trends and quantify the scale and dynamics of COVID-19 infection in a specific community, municipality, or area of service. This study investigates a six-month long SARS-CoV-2 RNA quantification in influent wastewater from four municipal wastewater treatment plants (WWTP) serving the Charlotte region of North Carolina (NC) using both RT-qPCR and RT-ddPCR platforms. Influent wastewater was analyzed for the nucleocapsid (N) genes N1 and N2. Both RT-qPCR and RT-ddPCR performed well for detection and quantification of SARS-CoV-2 using the N1 target, while for the N2 target RT-ddPCR was more sensitive. SARS-CoV-2 concentration ranged from 10^3^ to10^5^ copies/L for all four plants. Both RT-qPCR and RT-ddPCR showed a significant moderate to a strong positive correlation between SARS-CoV-2 concentrations and the 7-day rolling average of clinically reported COVID-19 cases using a lag that ranged from 7 to 12 days. A major finding of this study is that despite small differences, both RT-qPCR and RT-ddPCR performed well for tracking the SARS-CoV-2 virus across WWTP of a range of sizes and metropolitan service functions.

**Graphical Abstract:** 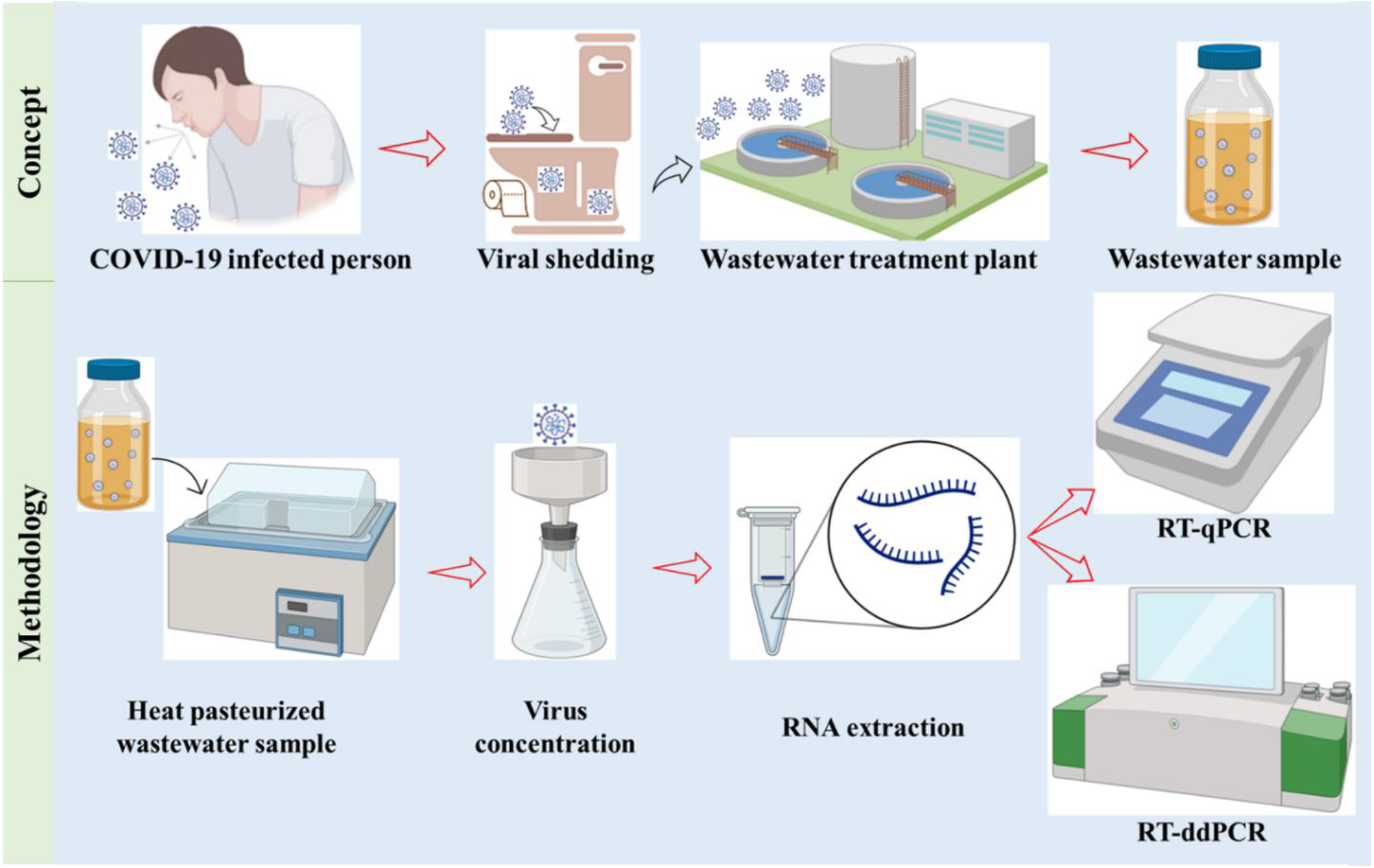

## 1. Introduction

The global pandemic “Coronavirus disease 2019 (COVID-19)”, as declared by the World Health Organization (WHO, 2020a), is caused by the virus given the name “Severe Acute Respiratory Syndrome Coronavirus 2” (SARS-CoV-2). The single-stranded ribonucleic acid (RNA) SARS-CoV-2 virus can infect individuals causing a range of symptoms, which can include life-threatening health complications on one end of the spectrum or a lack of symptoms (asymptomatic carriers). Interestingly, both symptomatic and asymptomatic individuals have the potential to spread the virus to others in the population (Bai et al., 2020). This makes tracking infected individuals and implementing appropriate preventative measures difficult.

During the onset of the COVID-19 pandemic, clinical testing was restricted primarily to individuals exhibiting life-threatening health complications owing to limited COVID-19 clinical testing kits (CDC, 2020). Thus, many asymptomatic and even symptomatic individuals were excluded from the COVID-19 case counts when public health decisions were made (Murakami et al., 2020) during the early stages of the pandemic. Although later stages of the pandemic have included testing of asymptomatic individuals, for either surveillance or screening, testing has been neither comprehensive nor representative. Therefore, clinical testing has been valuable for managing isolation and quarantine of individuals, but the pooling of clinical testing data has limited utility for understanding overall trends or inferring the prevalence of infection in entire communities/counties.

Monitoring of SARS-CoV-2 in wastewater influent from municipal wastewater treatment plants (WWTP) has been demonstrated to be a useful tool for predicting clinical outcomes for whole communities (Agrawal et al., 2021; Ahmed et al., 2021; Hillary et al., 2021; Saguti et al., 2021). Wastewater influent is an aggregate measure of the prevalence of infection in a community, particularly for viral, bacterial and protozoan pathogens that are carried in fecal material. SARS-CoV-2 RNA concentration in wastewater influent has not only been correlated with reported COVID-19 cases, but they have been predictive of the clinical testing outcomes in communities sometimes with as much as a 6 to14 day lead time (Kumar et al., 2021; Peccia et al., 2020). Monitoring of influent wastewater has revolutionized the tracking of pathogens in municipalities, communities, and even small-scale systems such as dormitories and workplaces. Monitoring of SARS-CoV-2 in wastewater influent includes virus being shed from symptomatic, clinically diagnosed, and asymptomatic individuals. This area of active research will yield beneficial information for guiding public health decisions.

WBE is a potential approach for understanding the proliferation of SARS-CoV-2 within a community as the viral RNA is shed by infected individuals into wastewater (Hasan et al., 2021; Hemalatha et al., 2021). Aoust et al. (2021) reported that the surges in SARS-CoV-2 RNA in wastewater were observed 48 h prior to clinical testing and 96 h prior to hospitalization. Wastewater sampling captures the community signal comprising both symptomatic and asymptomatic individuals (Bivins et al., 2020; Peccia et al., 2020), suggesting the value of WBE as an impartial surveillance system at a community level when making public health decisions. To date, numerous studies have documented the detection of SARS-CoV-2 in the influent of municipal wastewater i.e., Ahmed et al. (2020); Albastaki et al. (2021); Bertrand et al. (2021); Gonçalves et al. (2021); Kumar et al. (2020); to name a few but their study period ranged from only 15-30 days. Weidhaas et al. (2021) articulated the need for a meticulous WBE study for prolonged periods in localities with lower and higher COVID-19 cases to identify the relationship between concentrations of SARS-CoV-2 RNA in municipal wastewater and rates of COVID-19 cases in the corresponding communities.

This manuscript details a six-month long WBE study for the surveillance of SARS-CoV-2 in the influent municipal wastewater of Charlotte, North Carolina (NC). The number of clinical cases of COVID-19 in Mecklenburg County, where Charlotte is located, was highest among all the counties of NC. The most populous city in NC, Charlotte includes the Charlotte Douglas International Airport. By December 2020, the number of COVID-19 cases was reported to be greater than 65,000 in Mecklenburg County (North Carolina Department of Health and Human Services, NCDHHS). Fig. 1 shows Mecklenburg County where Charlotte is located to report the highest number of COVID-19 cases in NC. As of September 11, 2021, Mecklenburg County leads the state in total reported COVID-19 cases with 141,000.

**Fig. 1.**
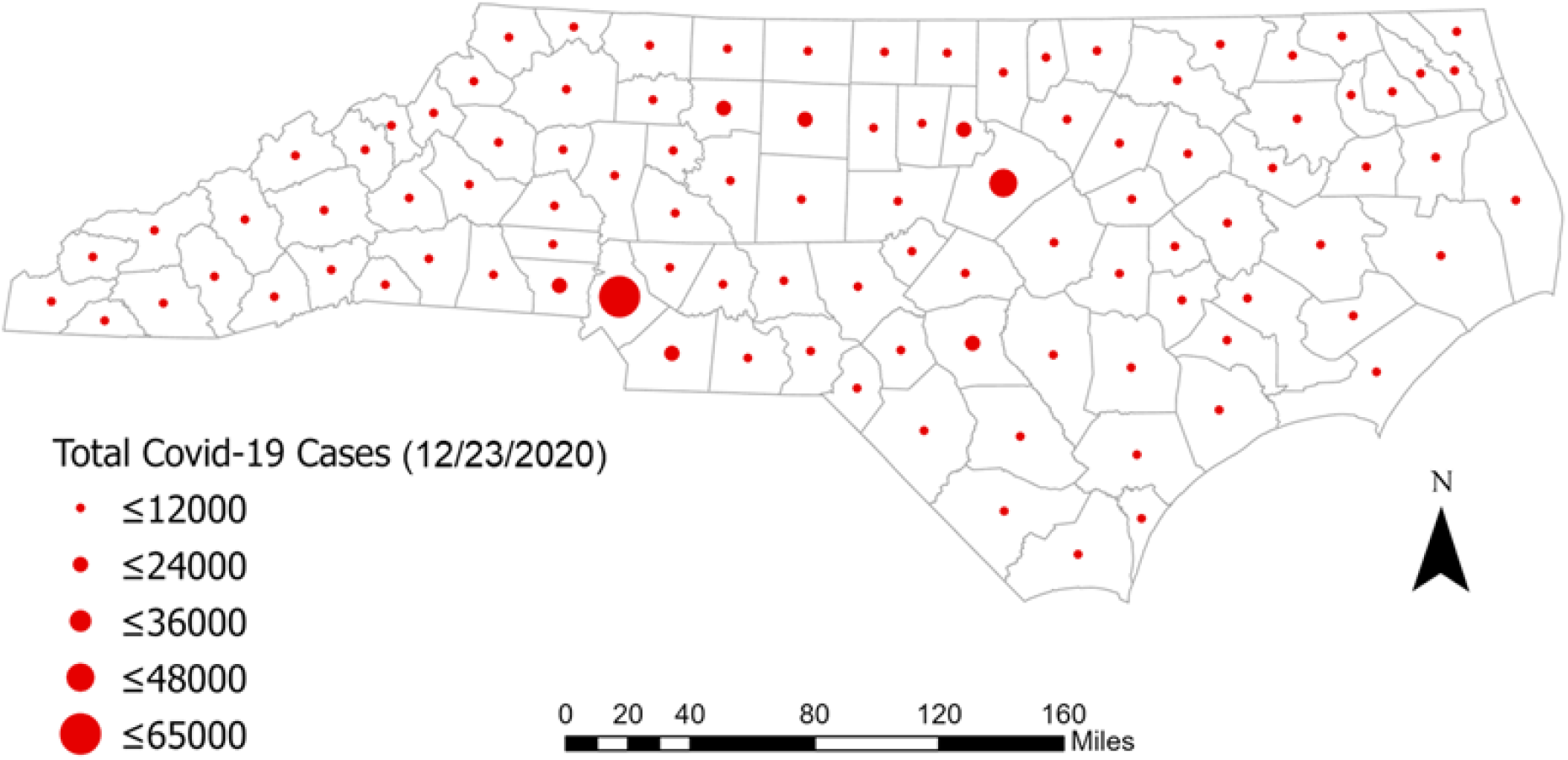
Map showing the total number of clinically reported COVID-19 cases, county-wise, in the state of NC for the duration of this study (Prepared by the software ArcGIS Pro).

To date, SARS-CoV-2 wastewater surveillance studies have mostly employed RT-qPCR for viral quantification (Ahmed, Angel, et al., 2020; Chik et al., 2021; Gerrity et al., 2021; Haramoto et al., 2020; Medema et al., 2020; Nemudryi et al., 2020; Peccia et al., 2020; Randazzo et al., 2020; Sherchan et al., 2020; Westhaus et al., 2021; Wurtzer et al., 2020; Zhao et al., 2021) rather than RT-ddPCR (Gonzalez et al., 2020; Gonzalez et al., 2021). Only a few research groups have used both RT-qPCR and RT-ddPCR quantification (Aoust, Graber, et al., 2021; Ciesielski et al., 2021; Dumke et al., 2021; Graham et al., 2021). The study conducted by both Graham et al. (2021) and Aoust et al. (2021) focussed on RT-qPCR and RT-ddPCR quantification for solids from WWTP, while Dumke et al. (2021) targeted E and S genes to quantify SARS-CoV-2 in wastewater. Ciesielski et al. (2021) performed an interlaboratory validation study of 60 samples comparing RT-qPCR to RT-ddPCR quantification. The aim of this study was to (a) compare the utilization of two different molecular quantification platforms to identify the changing aspects of SARS-CoV-2 viral concentration in the wastewater influent from four WWTP serving Charlotte, North Carolina (NC) for six months, and (b) to correlate wastewater concentration (quantified by both RT-qPCR and RT-ddPCR) with clinical surveillance data of SARS-CoV-2 infection.

## 2. Methodology

### 2.1 Sample collection

24-h flow-weighted composite samples of influent wastewater were initially collected every Wednesday starting on June 24, 2020, from four WWTP (A, B, C, and D) in Charlotte, North Carolina. Wastewater samples were collected in the morning between 7:00-8:45 am in sterile 1L Nalgene bottles. Following collection, the wastewater samples were heat pasteurized for 40 minutes at 75°C in abidance with the Institutional Biosafety Committee’s mandatory protocol for the protection of laboratory personnel (WHO, 2020b). Heat pasteurized duplicate samples from each WWTP were transported to the laboratory in coolers packed with ice. Deionized water in a 1 L Nalgene sample collection bottle was used as a field blank. The field blank was exposed to the same environment and transported to the laboratory in coolers packed with ice along with the wastewater samples. The collected samples were processed immediately after reaching the laboratory. A recent study conducted by Pecson et al. (2021) observed that SARS-CoV-2 quantitation was slightly higher in pasteurized samples after recovery correction. Sample collection increased to twice a week, on Monday and Wednesday during November and December 2020. Monday sampling represented the 24 h composite sample beginning on Sunday through Monday while Wednesday sampling represented the 24 h composite sample beginning on Tuesday through Wednesday. A total of 115 wastewater samples were collected during 31 sampling events. Data from two sampling events were not included in this reported dataset because the PBS blank demonstrated cross-contamination of the samples. The characteristics of each of the WWTP have been provided in Table1.

**Table 1:**
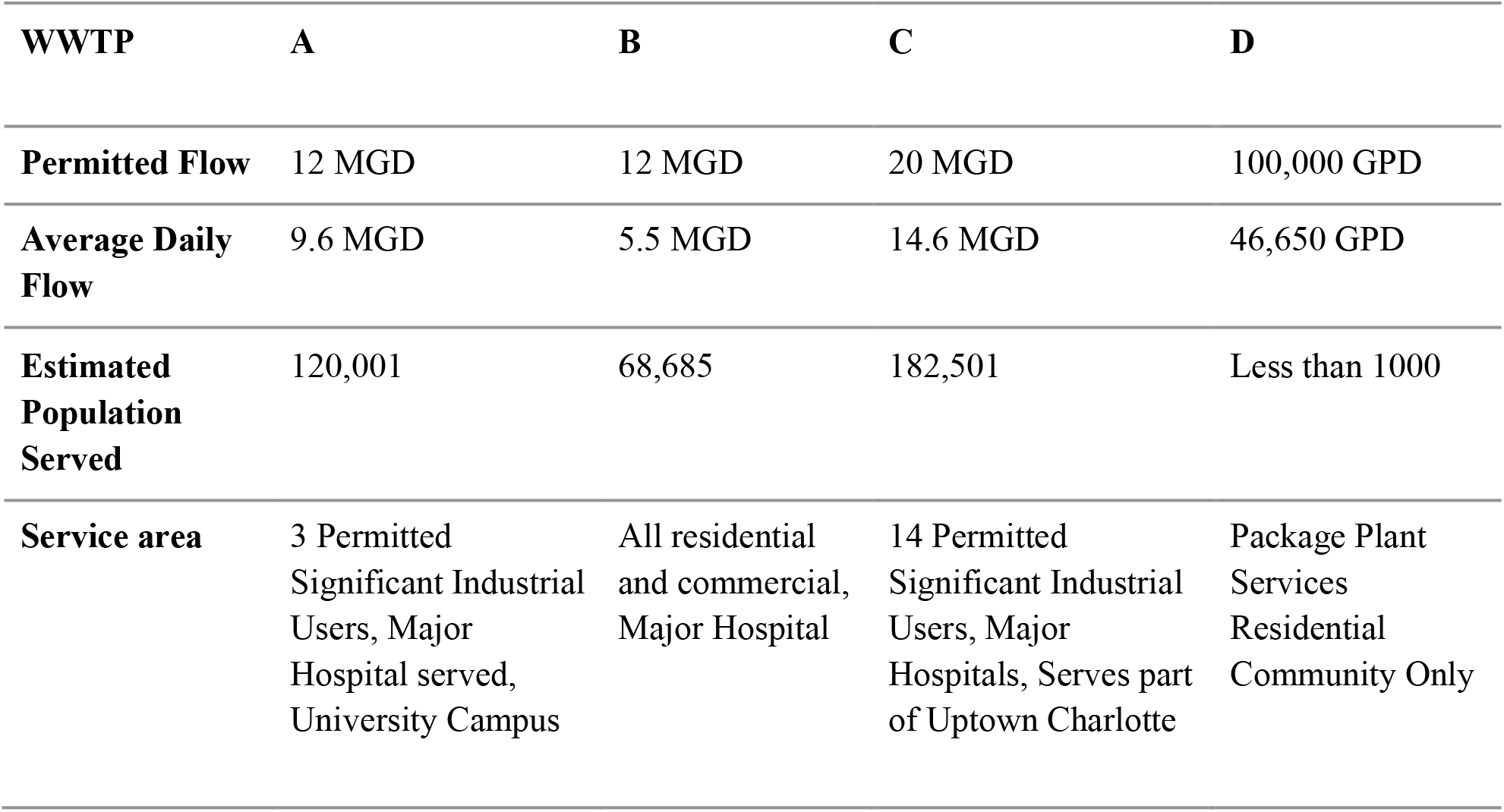
Wastewater Treatment Plant (WWTP) characteristics.

| WWTP | A | B | C | D |
| --- | --- | --- | --- | --- |
| Permitted Flow | 12 MGD | 12 MGD | 20 MGD | 100,000 GPD |
| Average Daily Flow | 9.6 MGD | 5.5 MGD | 14.6 MGD | 46,650 GPD |
| Estimated Population Served | 120,001 | 68,685 | 182,501 | Less than 1000 |
| Service area | 3 Permitted Significant Industrial Users, Major Hospital served, University Campus | All residential and commercial, Major Hospital | 14 Permitted Significant Industrial Users, Major Hospitals, Serves part of Uptown Charlotte | Package Plant Services Residential Community Only |

### 2.2 Sample concentration

6300 copies/μL of Bovine Coronavirus (BCoV, ValleyVet Supply, Marysville, KS) were spiked into the wastewater sample before concentration as overall process control. Wastewater samples were adjusted to a pH of 3.5-4 using 10M HCl, followed by the addition of 2.5 M MgCl_2_.6H_2_O to achieve a final concentration of 25 mM MgCl_2_.6H_2_O (Ahmed, Bertsch, et al., 2020; Cashdollar and Wymer, 2013). Using a disposable filter funnel fitted with a 47 mm dia. 0.45 μm type HA Filter (Millipore, Bedford MA), 20 mL of each wastewater sample was concentrated using a vacuum filtration manifold and was filtered to dryness. Negative process control or Method Blank (MB) consisting of 1X phosphate buffered saline (PBS) was filtered during each of the sample processing events using a new sterile filter funnel and type HA electronegative filter (Ciesielski et al., 2021). After wastewater concentration, the filter was placed in individual 2 mL microcentrifuge tubes. The process was repeated 8 times for each wastewater sample. One filter was used for Workflow 1, one for Workflow 2, (Fig. 2) and the others were archived at - 80°C for future analyses. For workflow 1 the filter was suspended in the AVL buffer for RNA extraction.

**Fig. 2.**
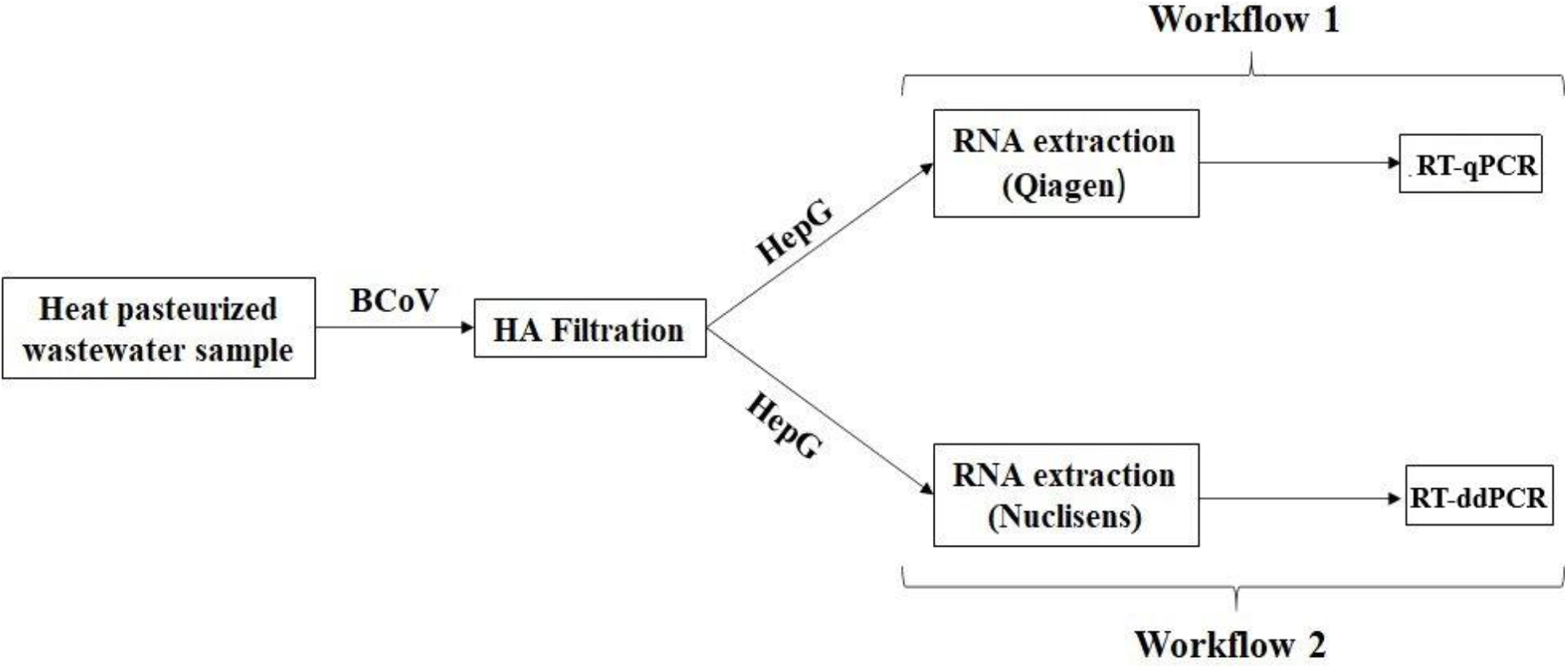
Showing the two different workflows performed to quantify SARS-CoV-2 in the influent wastewater.

### 2.3 Workflow 1

#### 2.3.1 Viral RNA extraction

The filters with concentrated samples were suspended in 1000 μL of AVL lysis buffer with carrier RNA and spiked with 15,600 copies of armored Hepatitis G (Hep G) (p/n 42024 Asuragen, Austin, TX). Samples were then vortexed and incubated at room temperature for 10 minutes to facilitate viral recovery from the filter surface (Gibas et al., 2021; Juel et al., 2021). QIAamp® Viral RNA Mini Kit (Qiagen, Germantown, Maryland, USA) was used following the manufacturer’s instructions for viral RNA extraction where the amount of the lysed sample was 200 μL with a final elution volume of 60 μL of viral RNA extract.

#### 2.3.2 Detection and quantification using RT-qPCR

Detection and quantification of SARS-CoV-2 viral RNA in wastewater were performed by one-step RT-qPCR on a CFX Connect thermocycler (Bio-Rad, Hercules, CA) utilizing the 2019-nCoV CDC RUO Kit (Integrated DNA Technologies) targeting the nucleocapsid genes (N1 and N2) (Table S1). The reaction mixture comprised a total volume of 20 μL containing 5 µL extracted RNA template, 10 µL iTaq universal probes reaction mix (Bio-Rad), 0.5 µL iScript reverse transcriptase (Bio-Rad), 1.5 µL (500 nM) primers along with a (125 nM) probe and 3 µL of nuclease-free water. The thermocycling conditions employed were 25°C for 2 min, 50°C for 15 min, 95°C for 2 min followed by 45 cycles of amplification including denaturation at 95 °C for 3 secs and extension at 55 °C for 30 secs (CDC RT-qPCR panel 2020). Synthetic, single-stranded SARS-CoV-2 RNA (Twist Bioscience, San Francisco, CA) was used as a positive control. No template control (NTC) in triplicate was included with every run, where the RNA template was replaced with nuclease-free water, to determine if the mastermix was contaminated and if there was non-specific amplification during the later amplification cycles. Each sample was analyzed in triplicate, including the positive control and NTC reactions on each RT-qPCR run. RT-qPCR runs were analyzed by Bio-Rad CFX Manager software version 3.1 (Bio-Rad Laboratories).

#### 2.3.3 Quality Control (QC) Parameters

Precise QC metrics were considered to assess the detection sensitivity of CDC recommended N1 and N2 assays for both workflow 1 (RT-qPCR) and workflow 2 (RT-ddPCR). QC was taken into consideration throughout the whole study to avoid ambiguous interpretation of the obtained results. The positive and negative controls used during each of the steps for both the workflows (1 and 2) were in accordance with MIQE (Bustin et al., 2009) and the digital MIQE (dMIQE Group, 2020) guidelines. The detailed quality control and the criteria for data evaluation implemented has been provided below;

##### 2.3.3.1 Process Control

BCoV was spiked into wastewater samples as a proxy for SARS CoV-2, which could be measured throughout the extraction and RT-qPCR process. 6300 copies of BCoV vaccine was spiked per mL of wastewater. The initial titer of BCoV vaccine was quantified by RT-ddPCR prior to spiking. The average BCoV recovery for each of the WWTP was observed to be 21-31%.

##### 2.3.3.2 Extraction control

15,600 copies of armored hepG were spiked into the lysis buffer before the RNA extraction process to check the quality of the extracted RNA. The initial concentration of the armored hepG was determined by ddPCR after heat treatment at 75° for 3 minutes to remove the protein coat surrounding the HepG RNA sequence. The average HepG recovery for each of the WWTP was observed to be 38-44%.

##### 2.3.3.3 Standard Curve

Single-stranded RNA from Twist Bioscience was extracted in the same manner as wastewater influent samples. The RNA standard was quantified using RT-ddPCR prior to extraction. 10-fold serial dilution was performed with the extracted RNA over four orders of magnitude for generating N1 and N2 standard curves. Detailed information has been provided in the supplementary file (Fig. S1). The amplification efficiency was 90% for both N1 and N2 assay with an R^2^ value of 0.998 and 0.997, respectively which was within the acceptable range as specified in MIQE guidelines (Bustin et al., 2009).

##### 2.3.3.4 Limit of Detection

To avoid false positives and provide precise quantification, the limit of detection (LoD) for the assay was determined by running an extended series of dilutions of the RNA based SARS-CoV-2 positive control (Twist Bioscience) in six replicates with as few as 1 copy/reaction (three-fold dilution series towards the lower end). The threshold cycle at which signals were observed for all the three replicates with a standard deviation less than 1 was considered to be the Cq of LoD (*Cq*_*LoD*_). Cq values of 37.07 and 37.78 for N1 and N2 assays, respectively were converted to copies per reaction using the equation (1) to get the LoD for the assay.

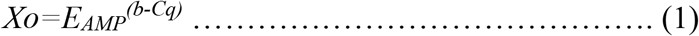

Where, *E*_*AMP*_ represents exponential amplification value of RT-qPCR assay, evaluated as *E*_*AMP*_= 10^−1/*m*^, *b* represents the intercept and *m* represents the slope. The LoD for workflow 1 was determined as 3000 copies/L of wastewater for both targets.

##### 2.3.3.5 Inhibition

The dilution method was used for the determination of the RT-qPCR inhibition (Graham et al., 2021). A dilution series of 1:1, 1:2, 1:5 and 1:10 was performed on a subset of samples (n=10) for assessing inhibition. If the diluted sample showed a more than 1 Cq difference between the actual and theoretically expected change in Cq, then the undiluted samples were considered inhibited. There was no inhibition observed for the N1 target but there was with N2. A dilution 1:2 was selected to continue inhibition testing as further dilution resulted in Cq values beyond LoD or as non-detectable and the quantification data was updated accordingly.

##### 2.3.3.6 Other Criteria for QC and data evaluation

- RNA extraction and master mix preparation for molecular quantification was conducted in two different biosafety cabinets in two separate laboratories next to each other to reduce contamination potential.
- RNA samples showing very poor overall recovery (below 2%) compared to the average recovery (23%) were re-extracted and re-quantified.
- Samples were considered positive when a minimum of two out of three replicates showed amplification above LoD for N1 and N2 assay.

### 2.4 Workflow 2

#### 2.4.1 Viral RNA extraction

Frozen filters containing the concentrated sample were shipped on dry ice and stored at -80° until analysis. The filter containing the concentrated sample was placed in 1mL of Nuclisens® easyMAG® Lysis Buffer (Biomerieux, Durham, NC) containing approximately 900 copies of armored HepG and incubated for a minimum of 10 minutes at room temperature. Lysis tubes were centrifuged for 1 minute at 13,000 x *g* and up to 950 μL of lysate transferred to a 96 well deep well plate (DWP). All samples, including controls, were extracted using NucliSens® EasyMAG reagents (Biomerieux, Durham, NC) on a KingFisher Flex (ThermoFisher, Waltham, MA) with a final elution volume of 100μL. KingFisher script is provided in the supplementary material (Table S7a).

#### 2.4.2 Detection and quantification using RT-ddPCR

RT-ddPCR was utilized to quantify SARS-CoV-2 RNA copies targeting N1 and N2, described previously (Table S1), and utilizing a two-step reverse transcription and RT-ddPCR. Purified RNA was reverse transcribed using Superscript VILO IV MM (ThermoFisher Waltham, MA.). Briefly, 50μL of the eluate was combined with 20μL 5X VILO IV MM, 1μL (160 copies) mouse lung RNA (p/n R1334152-50 BioChain Newark, CA) and 29μL of DEPC water for a total reaction volume of 100 μL (Table S7a). Reverse transcription was performed on a C1000 deep block thermal cycler (BioRad) with the following conditions: 25° for 10 minutes, 50° for 10 mins, and 85° for 5 minutes. 5 μL of cDNA was used for each RT-ddPCR reaction. A mastermix was created by the addition of forward and reverse primers (0.9µM final concentration) and for probes (0.25µM final concentration), 12.5µL of 2X Supermix for Probes (no dUTP, Bio-Rad), 5µL template, and nuclease-free water for a final volume of 25µL. A minimum of 4 no template controls (NTC), which substituted 5µL nuclease-free water for the template, were included in each run with every assay plate. Primers and probes were synthesized by LGC Biosearch Technologies (Novato, CA) except for Mouse ACTB exogenous control (Life Technologies ThermoFisher Scientific Waltham, MA). The concentration used in the assays is listed in S7b. Primers and probe sequences for the *gyr*A for inhibition control were kindly provided by John Griffith (SCCWRP) and have not been published. The inhibition probe was labeled with the HEX fluorophore and the RT-ddPCR assay was run as a duplex with all reactions performed in duplicate. Positive and negative controls were run on every assay plate. All assay conditions were previously optimized and established by the Noble Laboratory. Droplet generation was performed in accordance with manufacturer’s instructions, and then droplets were amplified in a C1000 thermal cycler with the following temperature profile: 10 min at 95°C for initial denaturation, 40 cycles of 94°C for 30 s, and 55°C for 60 s, followed by 98°C for 10 min, with a ramp rate of 2° per sec, then an indefinite hold at 12°. After RT-ddPCR cycling was complete, the plate was placed in a QX200 instrument (Bio-Rad) and droplets were analyzed according to the manufacturer’s instructions. Data acquisition and analysis were performed with QuantaSoft V1.74.0917 (Bio-Rad). The fluorescence amplitude threshold, distinguishing positive from negative droplets, was set manually by the analyst as the midpoint between the average baseline fluorescence amplitude of the positive and negative droplet cluster. The same threshold was applied to all the wells of one RT-ddPCR plate.

Measurement results of single RT-ddPCR wells were excluded on the basis of technical reasons in case that (i) the total number of accepted droplets was <10,000, or (ii) the average fluorescence amplitudes of positive or negative droplets were clearly different from those of the other wells on the plate. The numbers of positive and accepted droplets and concentration per μL were transferred to an in-house developed spreadsheet to calculate the copy number per filtered volume. Replicate wells were merged, and a sample was considered positive only if there were three or more positive droplets and each well contained a minimum of 10,000 droplets.

#### 2.4.3 Process Control

BCoV was spiked into wastewater samples as a proxy for SARS CoV-2, which could be measured throughout the extraction and RT-qPCR process. The copy number of BCoV was quantified by RT-ddPCR prior to spiking. The filter was extracted utilizing the same viral RNA extraction kit as the influent wastewater samples. About 38-44% average BCoV recovery was observed for each of the WWTP.

#### 2.4.4 Extraction control

Approximately 900 copies of armored HepG were spiked into the Lysis Buffer before the RNA extraction process to monitor the quality of the extracted RNA. Negative extraction controls (NECs) were included to verify the absence of cross-contamination and consisted of a blank HA filter processed under the same conditions as the other samples. The initial concentration of the armored HepG was determined by RT-ddPCR after heat treatment at 75° for 3 minutes to remove the protein coat surrounding the HepG RNA sequence. The average HepG recovery for all the WWTP was found to be 17.3 - 29.8%.

#### 2.4.5 Inhibition control

PCR inhibition was measured by the addition of a halophilic archaeon containing 160 copies of the gyrA gene into the mastermix. The halophiles had been cultured, aliquots frozen at -20°, and the concentration determined independently prior to the sample analysis. Inhibition was measured by the addition of exogenous cells and a sample was deemed inhibited if the difference of the expected versus the actual concentration differed by greater than 0.5 log (Table S6).

#### 2.4.6 Reverse transcription (RT) efficiency control

162 copies of mouse lung total RNA were spiked into the reverse transcription master mix and the recovery was measured using a mouse ACTB assay (Life Technologies). Recovery was measured by dividing the concentration of the unknown sample by the negative extraction control and multiplying by 100 (Table S7b).

#### 2.4.7 N1 and N2 Standard

Armored RNA Quant SARS-CoV-2 control, which encapsulates the in vitro transcribed RNA template in a protective protein coat and targets the SARS-CoV-2 viral nucleocapsid (N) region, was used as a positive control and run in duplicate for every set of reactions targeting N1 and N2.

#### 2.4.8 Limit of Detection, Limit of Quantification, and Limit of Blank

For the determination of LoD using RT-ddPCR, the Limit of Blank (LoB) was elucidated from eight replicates of negative matrix samples derived from influent collected at multiple WWTP throughout eastern NC. The LOB was calculated as the mean concentration of all sixty-four replicates and the LOD was then calculated as two standard deviations beyond the defined LOB (Hayden et al., 2013). The LOQ was determined to be never less than 3 positive droplets no matter the number of merged wells, which for this study was two, resulting in 10 μL or 10% of the RNA eluate and is equal to a concentration of 10 copies. The detailed LOB, LOD, and LOQ for N1 and N2 gene targets for RT-ddPCR has been provided in Table 2.

**Table 2:**
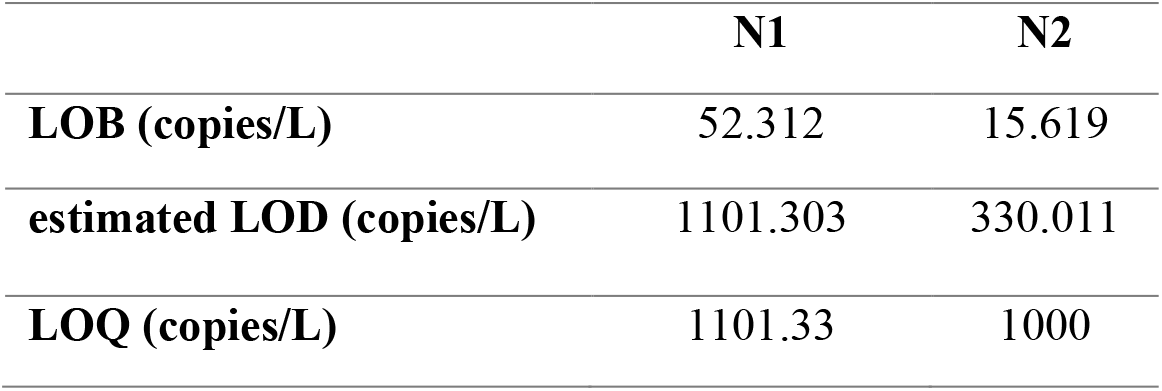
LOB, LOD, and LOQ for N1 and N2 gene targets for RT-ddPCR.

### 2.5 Recovery efficiency of BCoV and HepG

The following formula was utilized for both workflow 1 and 2 to determine the recovery efficiency of BCoV and HepG;

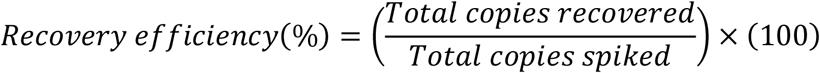

The average BCoV and HepG recovery efficiency for workflows 1 and 2 are provided in Supplementary Tables S2 and S3.

### 2.6 Epidemiological data

The North Carolina Department of Health and Human Services (NC DHHS) published the cumulative confirmed COVID-19 cases by 5-digit zip code as an online map (https://nc.maps.arcgis.com/home/item.html?id=52f127a0767149ec984e91fcc06b06cb#overview). The map was typically updated daily, overwriting the previous day’s count. We obtained a daily time series of cumulative cases from the WRAL online repository (https://github.com/wraldata/nc-covid-data/tree/master/zip_level_data/time_series_data/csv), which extracted and archived the NC DHHS published case reports each day. Missing counts in the WRAL archive were filled with the cumulative cases reported for the same date that we had manually archived from the NC DHHS COVID-19 dashboard for a subset of dates (https://covid19.ncdhhs.gov/dashboard/data-behind-dashboards). We calculated daily incident cases as the difference between the current and previous day’s reported cumulative cases, carrying the last non-missing value forward as necessary.

Zip code and sewershed boundaries do not typically align (Fig.3). Daily case counts for each sewershed were represented by the sum of all cases in each zip code that substantially overlapped the sewershed boundary, defined as >50% of the zip code area within the sewershed or >50% of the sewershed area within the zip code. We used the 2019 American Community Survey (ACS) 5-year block group population estimates to estimate the population served by each sewershed.

**Fig. 3.**
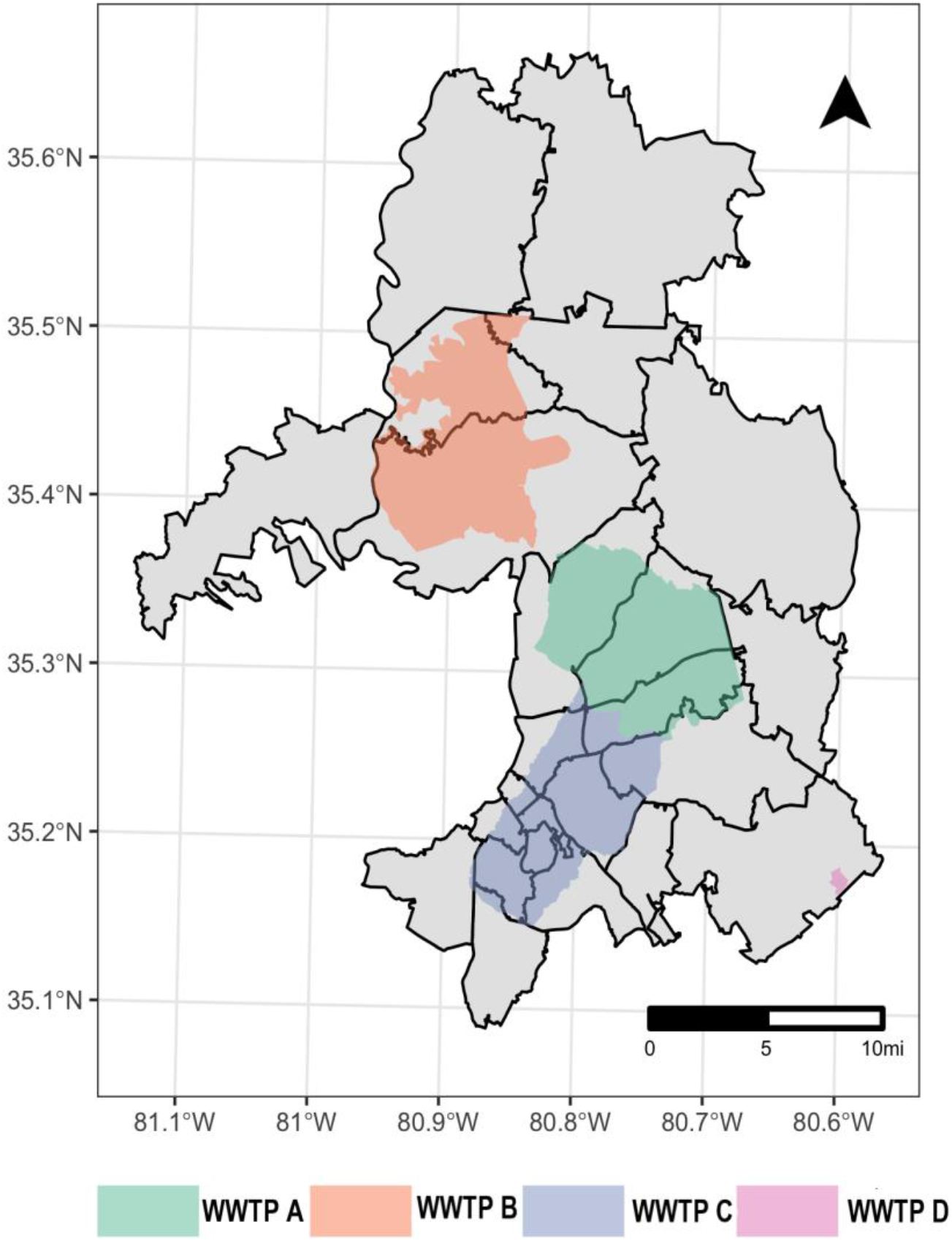
Map showing the four sewershed location and the overlapping zip codes of Charlotte, NC.

### 2.7 Statistical analysis

Percent agreement statistics and Cohen’s Kappa coefficient was used to determine the agreement of SARS-CoV-2 positivity and negativity results between the RT-qPCR and RT-ddPCR (McHugh, 2012; Obermeier et al., 2016). The strength of the agreement is interpreted based on the Kappa value (k): there is no agreement if k ≤ 0, slight agreement if k=0.01-0.20, fair if k= 0.21-0.40, moderate if k= 0.41-0.60, substantial if k=0.61-0.80, and nearly a perfect agreement if k=0.81-1.00 (McHugh, 2012). Spearman’s rank correlation test was performed to determine the correlation of the SARS-CoV-2 concentration in influent wastewater with the averaged clinical (7-day moving average) COVID-19 cases. The correlation between the viral RNA signal and incident clinical cases, offset for 1 to 14 days (taking the wastewater influent collection date as the reference), was also computed for determining whether the influent wastewater SARS-CoV-2 RNA signal may serve as a leading indicator of the reported clinical cases. Case offset times exhibiting higher correlation with a p-value less than 0.05 were considered to be the probable lag time window.

## 3. Results and discussion

### 3.1 RT-qPCR vs RT-ddPCR platform

In this study, the utility of two different molecular platforms (RT-qPCR and RT-ddPCR) were compared to check SARS-CoV-2 detection frequency and concentration in the municipal influent wastewater.

#### 3.1.1 Detection frequency and trends

The detection frequency and trend of SARS-CoV-2 RNA in the municipal influent wastewater of Charlotte was observed by both RT-qPCR and RT-ddPCR using N1 and N2 targets. From the very first sampling event SARS-CoV-2 RNA was detected in the municipal wastewater influent samples of all the four WWTP throughout the six-month course (Fig. 4a). RT-qPCR detected a higher percentage of SARS-CoV-2 positives using the N1 target compared to N2 target (Table 3). About 27.83% of samples detected positive with the N1 target did not show any signal with the N2 target. In addition, the N2 assay showed inhibition while N1 did not (Table S4 and S5). On the other hand, RT-ddPCR performed well in detecting SARS-CoV-2 using both N1 and N2 targets, though the N2 target was quantified in a higher percentage (36-48%) of samples (Table 3). When comparing the molecular platform, RT-ddPCR showed more sensitivity than RT-qPCR in quantifying SARS-CoV-2 RNA in wastewater samples. However, SARS-CoV-2 RNA was quantified more readily using the N1 target across all samples using both platforms. As such, downstream analysis was conducted only using N1 data. SARS-CoV-2 positivity agreement between the two molecular platforms was 74.4% while the negative agreement was 52.6%. The overall percent agreement was 71% with the Cohen’s Kappa coefficient (k) of 0.21. Ciesielski et al. (2021) also found a similar agreement with a k value of 0.31 when comparing detection performance between RT-qPCR and RT-ddPCR. Other researchers compared RT-ddPCR and RT-qPCR and observed the former one to be more sensitive in the detection of low viral titer but they have mainly focused on N1 target only (Gonzalez et al., 2021).

**Table 3:** Detection frequency of N1 and N2 gene

| WWTP | RT-qPCR |  | RT-ddPCR |  |
| --- | --- | --- | --- | --- |
|  | N1 (%) | N2 (%) | N1 (%) | N2 (%) |
| A | 82.14 | 50 | 77.8 | 96.3 |
| B | 82.75 | 48.3 | 67.9 | 78.6 |
| C | 93.1 | 51.72 | 82.76 | 86.2 |
| D | 55 | 10 | 73.68 | 57.9 |

**Fig. 4.**
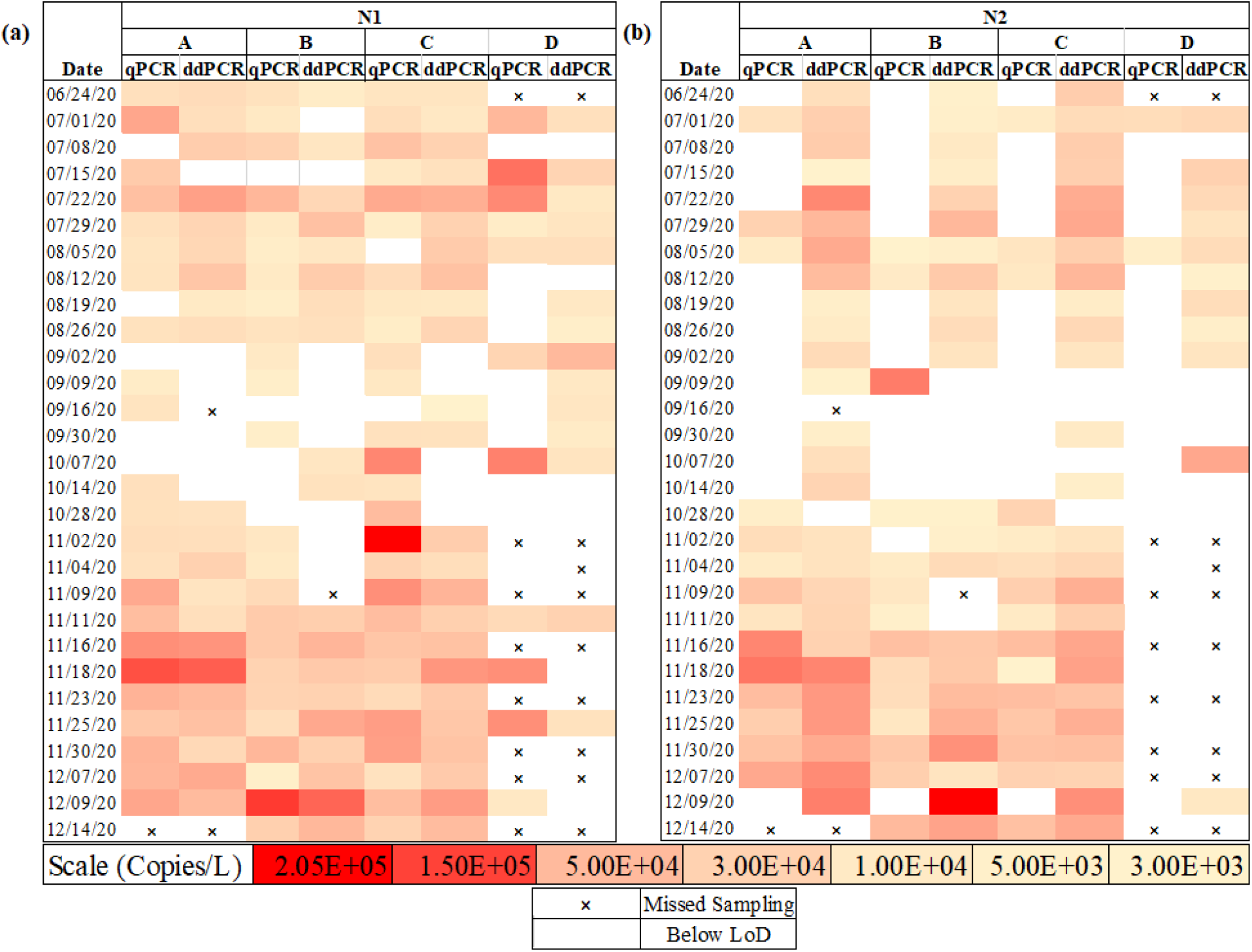
Heat map of concentrations of (a) N1 and (b) N2 targets to evaluate SARS-CoV-2 prevalence at WWTP A, B, C and D using RT-qPCR and RT-ddPCR. The symbol “×” indicates a missed sampling event and the uncolored blank spaces indicate a sample that was below the limit of detection (LoD).

#### 3.1.2 Quantitative relationship

The overall quantitative data generated using both RT-qPCR and RT-ddPCR for the WWTP A, B, C was positively correlated (ρ=0.569, p<0.0001) with statistical significance. The agreement between the platforms is shown using a range of colors corresponding to concentrations between 3.00E+03 copies/L and 2.05E+05 (Fig.4). RT-qPCR and RT-ddPCR generated similar SARS-CoV-2 RNA concentration data across the duration of the study which is indicated by the consistency between colors for both platforms on any given collection date. However, the quantitative data of WWTP D was highly variable and not significantly correlated (ρ=-0.047, p=0.91) which could be attributed to the fact that WWTP D is smaller in size and serves a smaller population compared to the other WWTP of Charlotte, NC. For most of the samples in this study, SARS-CoV-2 viral concentrations were in the range of 10^3^-10^5^ copies/L for both RT-ddPCR and RT-qPCR. These SARS-CoV-2 concentrations are consistent with previous studies conducted by Sherchan et al. (2020) and Gonzalez et al. (2020) in wastewater throughout Louisiana and Southeastern Virginia, respectively. The highest peak value of SARS-CoV-2 concentration in the influent wastewater for the WWTP A, B and C was observed to be around 1.15×10^5^-1.96×10^5^ copies/L by both RT-qPCR and RT-ddPCR. Also, the concentration of the 88% quantified samples were within 0.5 log variation resulting in a percentage difference within 12.5%. Miyani et al. (2020) also reported the highest SARS-CoV-2 concentration in the influent wastewater of Michigan to be within the range of 2×10^5^ copies/L. It is interesting to note that the highest viral quantification for both RT-ddPCR and RT-qPCR was observed by the end of November for WWTP A, B and C. Similar shades of colors (Fig. 4) were witnessed by the end of November indicating that the quantification by both RT-ddPCR and RT-qPCR were in agreement.

### 3.2 COVID-19 clinical cases and SARS-CoV-2 concentration in wastewater influent

The concentration of SARS-CoV-2 in the municipal influent wastewater was correlated with the clinically reported COVID-19 case numbers for Mecklenburg County, Charlotte, NC. The SARS-CoV-2 concentration quantified by both RT-qPCR and RT-ddPCR in the influent municipal wastewater of Charlotte for all the WWTP were plotted against the clinically reported 7-day average COVID-19 cases for zip codes served by each plant (Fig.5). From Fig. 5a, 5b and 5c it is evident that the trends of reported COVID-19 cases match with the influent wastewater concentration quantified by both RT-qPCR and RT-ddPCR. The influent wastewater concentration and the reported COVID-19 cases trends was a perfect match for WWTP A followed by WWTP C and B. For each WWTP, there was an increase during the summer months followed by a drop in both reported COVID-19 cases as well as influent wastewater concentration and then again, an increase was witnessed during the winter. In Charlotte, NC zip codes served by plants A and C mostly contributed to the increase in COVID-19 cases followed by WWTP B. Spearman rank correlation determined that there was a significant, moderate to strong, and positive correlation observed between SARS-CoV-2 RNA in influent wastewater and 7-day average COVID-19 cases throughout the entirety of the six-month period. This correlation became more robust when clinically reported COVID-19 cases were lagged in against the influent wastewater SARS-CoV-2 data. With RT-qPCR, the influent wastewater SARS-CoV-2 viral RNA data was likely to lead by 11 days (ρ=0.92, p<0.001), 10 days (ρ=0.81, p<0.001) and 5 days (ρ=0.61, p<0.001) for WWTP A, B, and C, respectively while using RT-ddPCR, the lead time was 12 days (ρ=0.67, p=0.001), 7 days (ρ=0.72, p<0.001) and 10 days (ρ=0.50, p<0.02) respectively. The lead time may vary depending on the sewershed pattern, the geospatial pattern of the population served for a WWTP, available testing facility and difference in the clinical sample collection date and result published to date (Bibby et al., 2021; Olesen et al., 2021). Even if the influent wastewater concentration data provided a predictive lead to the reported COVID-19 cases, it is interesting to note that the trend of the raw SARS-CoV-2 concentration data generated from the influent wastewater is similar to the reported COVID-19 cases. Additionally, SARS-CoV-2 concentration upsurge as quantified by both RT-qPCR and RT-ddPCR at a certain WWTP and decrease in another WWTP suggested that WBE provided us with the specific location where individuals are most or least infected than just the copies/L. Hasan et al. (2021) has also reported a similar observation where they have suggested the significant potential of WBE in monitoring upsurge or decline in COVID-19 positive case counts for a specific geographical location.

**Fig. 5.**
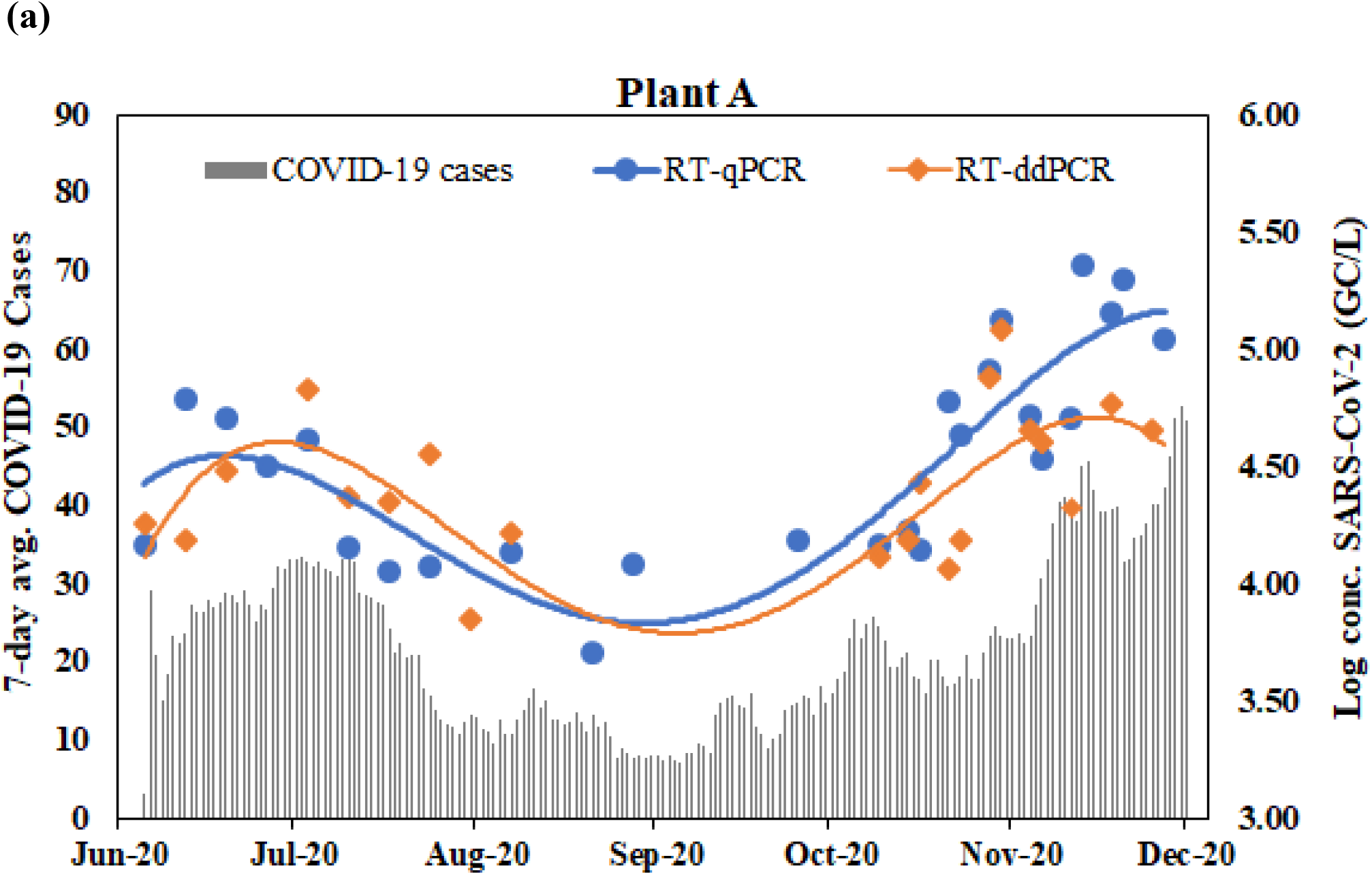
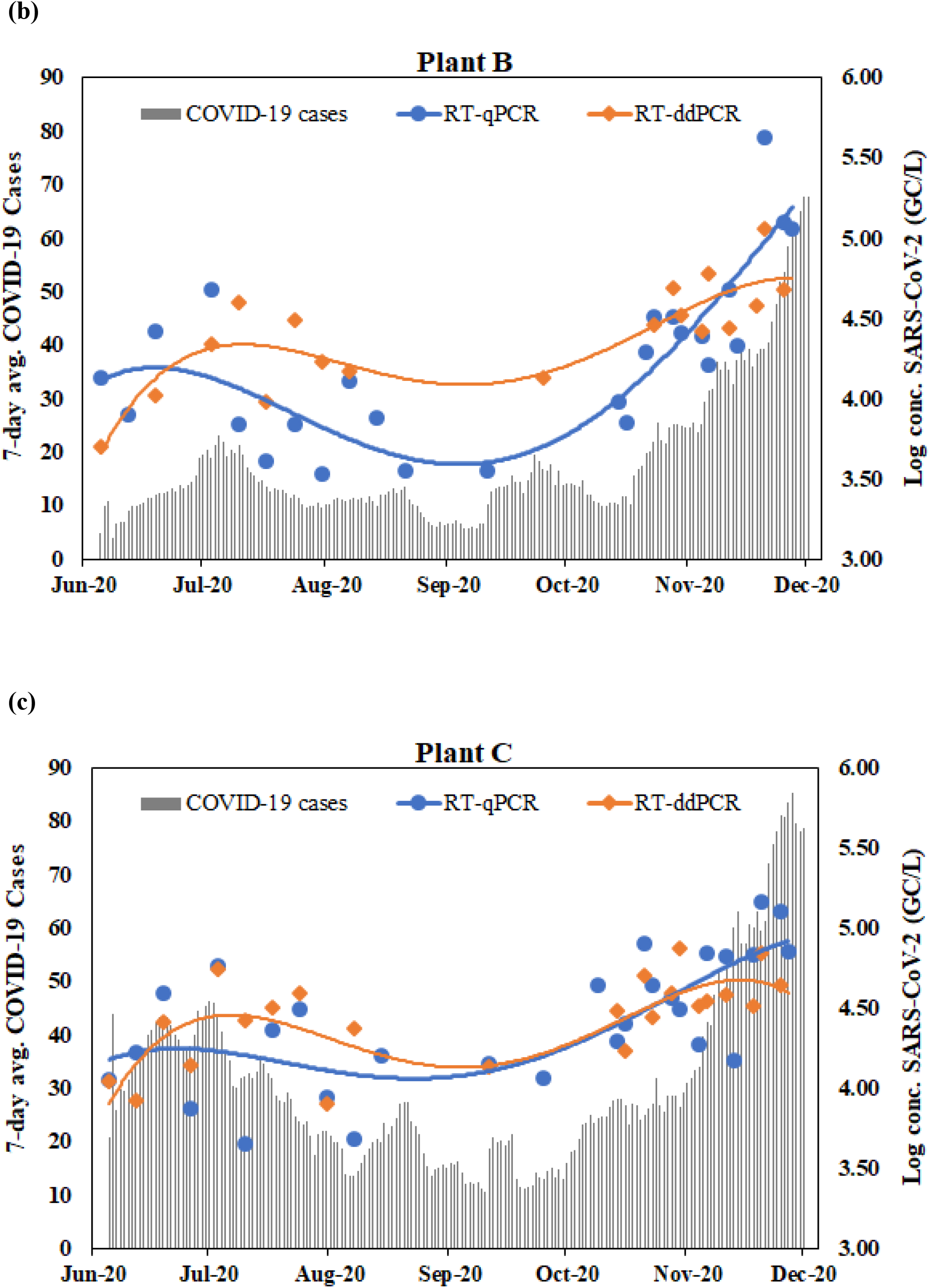
SARS-CoV-2 concentration (N1 target) for workflow 1 and 2 quantified by RT-qPCR and RT-ddPCR in the influent wastewater of (a) WWTP A, (b) WWTP B, and (c) WWTP C plotted against the 7-day average cases of each zipcode served by each WWTP. Quadratic polynomial trendline was used for the best fitted curve.

## 4. Conclusion

This long-term monitoring study of WWTP in the Charlotte Metropolitan area has demonstrated that wastewater-based monitoring for the N1 target can be successfully carried out using either RT-qPCR or RT-ddPCR. Different molecular platforms didn’t affect overall SARS-CoV-2 quantification in the influent wastewater and showed a good agreement with a variation of less than 12.5% for most of the samples. Depending on the WWTP, the Spearman rank correlation showed a moderate to a strong positive correlation between the influent wastewater SARS-CoV-2 viral signal and 7-day averaged reported COVID-19 cases. Importantly, influent wastewater SARS-CoV-2 RNA signal strength was leading the reported clinical COVID-19 cases by 5 to 12 days based on the WWTP, which is advantageous to monitor the COVID-19 outbreak in the community. Irrespective of the molecular platform used for the detection and quantification of SARS-CoV-2 in influent wastewater, it is important to incorporate all the QA/QC measures including implementation of appropriate external controls to obtain accurate and comparable results.

## Data Availability

Data is provided in the supplementary file and also available from the authors upon request

## Acknowledgment

This work was supported by North Carolina Policy Collaboratory. The authors acknowledge the support from the NC WW PATH team for early discussion and protocol sharing that was leveraged in this study. The authors would like to thank the Charlotte Water team including the wastewater treatment plant managers and operators for their support on wastewater sampling. The authors are grateful to Stacie Reckling (NC DHHS) and Steven Berkowitz (NC DHHS) for help with sewershed boundaries and other site related logistics. The authors are also grateful to Vivek Francis Pulikkal for supporting sample collection and preparation of the NC map using ArcGIS Pro software and Sol Park for helping with initial sample collection.

## References

Agrawal, S., Orschler, L., & Lackner, S. (2021). Long-term monitoring of SARS-CoV-2 RNA in wastewater of the Frankfurt metropolitan area in Southern Germany. Scientific Reports, 11(1), 5372. https://doi.org/10.1038/s41598-021-84914-2

Ahmed, W., Angel, N., Edson, J., Bibby, K., Bivins, A., O’Brien, J. W., Choi, P. M., Kitajima, M., Simpson, S. L., Li, J., Tscharke, B., Verhagen, R., Smith, W. J. M., Zaugg, J., Dierens, L., Hugenholtz, P., Thomas, K. V., & Mueller, J. F. (2020). First confirmed detection of SARS-CoV-2 in untreated wastewater in Australia: A proof of concept for the wastewater surveillance of COVID-19 in the community. Science of the Total Environment, 728, 138764. https://doi.org/10.1016/j.scitotenv.2020.138764

Ahmed, W., Bertsch, P. M., Bibby, K., Haramoto, E., Hewitt, J., Huygens, F., Gyawali, P., Korajkic, A., Riddell, S., Sherchan, S. P., Simpson, S. L., Sirikanchana, K., Symonds, E. M., Verhagen, R., Vasan, S. S., Kitajima, M., & Bivins, A. (2020). Decay of SARS-CoV-2 and surrogate murine hepatitis virus RNA in untreated wastewater to inform application in wastewater-based epidemiology. Environmental Research, 191(August), 110092. https://doi.org/10.1016/j.envres.2020.110092

Ahmed, W., Tscharke, B., Bertsch, P. M., Bibby, K., Bivins, A., Choi, P., Clarke, L., Dwyer, J., Edson, J., Nguyen, T. M. H., O’Brien, J. W., Simpson, S. L., Sherman, P., Thomas, K. V., Verhagen, R., Zaugg, J., & Mueller, J. F. (2021). SARS-CoV-2 RNA monitoring in wastewater as a potential early warning system for COVID-19 transmission in the community: A temporal case study. Science of the Total Environment, 761, 144216. https://doi.org/10.1016/j.scitotenv.2020.144216

Albastaki, A., Naji, M., Lootah, R., Almeheiri, R., Almulla, H., Almarri, I., Alreyami, A., Aden, A., & Alghafri, R. (2021). First confirmed detection of SARS-COV-2 in untreated municipal and aircraft wastewater in Dubai, UAE: The use of wastewater based epidemiology as an early warning tool to monitor the prevalence of COVID-19. Science of the Total Environment, 760, 143350. https://doi.org/10.1016/j.scitotenv.2020.143350

Aoust, P. M. D., Graber, T. E., Mercier, E., Montpetit, D., Alexandrov, I., Tariq, A., Mayne, J., Zhang, X., Alain, T., Servos, M. R., Srikanthan, N., Mackenzie, M., Figeys, D., Manuel, D., Jüni, P., Mackenzie, A. E., & Delatolla, R. (2021). Science of the Total Environment Catching a resurgence : Increase in SARS-CoV-2 viral RNA identi fied in wastewater 48 h before COVID-19 clinical tests and 96 h before hospitalizations. 770. https://doi.org/10.1016/j.scitotenv.2021.145319

Aoust, P. M. D., Mercier, E., Montpetit, D., Jia, J., Alexandrov, I., Neault, N., Tariq, A., Mayne, J., Zhang, X., Alain, T., Langlois, M., Servos, M. R., Mackenzie, M., Figeys, D., Mackenzie, A. E., Graber, T. E., & Delatolla, R. (2021). Quantitative analysis of SARS-CoV-2 RNA from wastewater solids in communities with low COVID-19 incidence and prevalence. Water Research, 188, 116560. https://doi.org/10.1016/j.watres.2020.116560

Bertrand, I., Challant, J., Mathieu, L., & Gantzer, C. (2021). International Journal of Hygiene and Environmental Health Epidemiological surveillance of SARS-CoV-2 by genome quantification in wastewater applied to a city in the northeast of France : Comparison of ultrafiltration-and protein precipitation-based metho. 233(January). https://doi.org/10.1016/j.ijheh.2021.113692

Bibby, K., Bivins, A., Wu, Z., & North, D. (2021). Making waves: Plausible lead time for wastewater based epidemiology as an early warning system for COVID-19. Water Research, 202(July), 117438. https://doi.org/10.1016/j.watres.2021.117438

Bivins, A., Greaves, J., Fischer, R., Yinda, K. C., Ahmed, W., Kitajima, M., Munster, V. J., & Bibby, K. (2020). Persistence of SARS-CoV - 2 in Water and Wastewater. https://doi.org/10.1021/acs.estlett.0c00730

Bustin, S. A., Benes, V., Garson, J. A., Hellemans, J., Huggett, J., Kubista, M., Mueller, R., Nolan, T., Pfaffl, M. W., & Shipley, G. L. (2009). The MIQE Guidelines : M inimum I nformation for Publication of Q uantitative Real-Time PCR E xperiments SUMMARY : 622, 611–622. https://doi.org/10.1373/clinchem.2008.112797

Cashdollar, J. L., & Wymer, L. (2013). Methods for primary concentration of viruses from water samples : a review and meta-analysis of recent studies. 1–11. https://doi.org/10.1111/jam.12143

CDC. (2020). 03/10/2020: Lab Advisory: Updated Guidance on Testing Persons for Coronavirus Disease 2019 (COVID-19). https://www.cdc.gov/csels/dls/locs/2020/updated_guidance_on_testing_persons_for_covid-19.html

Chik, A. H. S., Glier, M. B., Servos, M., Mangat, C. S., Pang, X. L., Qiu, Y., D’Aoust, P. M., Burnet, J. B., Delatolla, R., Dorner, S., Geng, Q., Giesy, J. P., McKay, R. M., Mulvey, M. R., Prystajecky, N., Srikanthan, N., Xie, Y., Conant, B., & Hrudey, S. E. (2021). Comparison of approaches to quantify SARS-CoV-2 in wastewater using RT-qPCR: Results and implications from a collaborative inter-laboratory study in Canada. Journal of Environmental Sciences (China), 107, 218–229. https://doi.org/10.1016/j.jes.2021.01.029

Ciesielski, M., Blackwood, D., Clerkin, T., Gonzalez, R., Thompson, H., Larson, A., & Noble, R. (2021). Assessing sensitivity and reproducibility of RT-ddPCR and RT-qPCR for the quantification of SARS-CoV-2 in wastewater. Journal of Virological Methods, July, 114230. https://doi.org/10.1016/j.jviromet.2021.114230

Dumke, R., De, M., Barron, C., Oertel, R., Helm, B., Kallies, R., Berendonk, T. U., & Dalpke, A. (2021). Evaluation of Two Methods to Concentrate SARS-CoV-2 from Untreated Wastewater. 1–7.

Gerrity, D., Papp, K., Stoker, M., Sims, A., & Frehner, W. (2021). Early-pandemic wastewater surveillance of SARS-CoV-2 in Southern Nevada: Methodology, occurrence, and incidence/prevalence considerations. Water Research X, 10, 100086. https://doi.org/10.1016/j.wroa.2020.100086

Gibas, C., Lambirth, K., Mittal, N., Islam, A., Bharati, V., Roppolo, L., Hinton, K., Lontai, J., Stark, N., Young, I., Quach, C., Russ, M., Kauer, J., Nicolosi, B., Chen, D., Akella, S., Tang, W., Schlueter, J., & Munir, M. (2021). Science of the Total Environment Implementing building-level SARS-CoV-2 wastewater surveillance on a university campus. Science of the Total Environment, 782, 146749. https://doi.org/10.1016/j.scitotenv.2021.146749

Gonçalves, J., Koritnik, T., Mio, V., Trkov, M., Bolje, M., Prosenc, K., Kotar, T., & Paragi, M. (2021). Science of the Total Environment Detection of SARS-CoV-2 RNA in hospital wastewater from a low COVID-19 disease prevalence area. 755, 4–10. https://doi.org/10.1016/j.scitotenv.2020.143226

Gonzalez, R. A., Larson, A., Thompson, H., Carter, E., & Cassi, X. F. (2021). Redesigning SARS-CoV-2 clinical RT-qPCR assays for wastewater RT-ddPCR. MedRxiv, 2021.03.02.21252754. https://doi.org/10.1101/2021.03.02.21252754

Gonzalez, R., Curtis, K., Bivins, A., Bibby, K., Weir, M. H., Yetka, K., Thompson, H., Keeling, D., Mitchell, J., & Gonzalez, D. (2020). COVID-19 surveillance in Southeastern Virginia using wastewater-based epidemiology. Water Research, 186, 116296. https://doi.org/10.1016/j.watres.2020.116296

Graham, K. E., Loeb, S. K., Wolfe, M. K., Catoe, D., Sinnott-armstrong, N., Kim, S., Yamahara, K. M., Sassoubre, L. M., Grijalva, L. M. M., Roldan-hernandez, L., Langenfeld, K., Wigginton, K. R., & Boehm, A. B. (2021). SARS-CoV-2 RNA in Wastewater Settled Solids Is Associated with COVID-19 Cases in a Large Urban Sewershed. Environmental Science & Technology. https://doi.org/10.1021/acs.est.0c06191

Haramoto, E., Malla, B., Thakali, O., & Kitajima, M. (2020). Science of the Total Environment First environmental surveillance for the presence of SARS-CoV-2 RNA in wastewater and river water in Japan. Science of the Total Environment, 737, 140405. https://doi.org/10.1016/j.scitotenv.2020.140405

Hasan, S. W., Ibrahim, Y., Daou, M., Kannout, H., Jan, N., Lopes, A., Alsafar, H., & Yousef, A. F. (2021). Science of the Total Environment Detection and quantification of SARS-CoV-2 RNA in wastewater and treated ef fl uents : Surveillance of COVID-19 epidemic in the United Arab Emirates. Science of the Total Environment, 764, 142929. https://doi.org/10.1016/j.scitotenv.2020.142929

Hayden, R. T., Gu, Z., Ingersoll, J., Abdul-Ali, D., Shi, L., Pounds, S., & Caliendo, A. M. (2013). Comparison of droplet digital PCR to real-time PCR for quantitative detection of cytomegalovirus. Journal of Clinical Microbiology, 51(2), 540–546. https://doi.org/10.1128/JCM.02620-12

Hemalatha, M., Kiran, U., Kumar, S., Kopperi, H., Gokulan, C. G., Mohan, S. V., & Mishra, R. K. (2021). Science of the Total Environment Surveillance of SARS-CoV-2 spread using wastewater-based epidemiology : Comprehensive study. Science of the Total Environment, 768, 144704. https://doi.org/10.1016/j.scitotenv.2020.144704

Hillary, L. S., Farkas, K., Maher, K. H., Lucaci, A., Thorpe, J., Distaso, M. A., Gaze, W. H., Paterson, S., Burke, T., Connor, T. R., McDonald, J. E., Malham, S. K., & Jones, D. L. (2021). Monitoring SARS-CoV-2 in municipal wastewater to evaluate the success of lockdown measures for controlling COVID-19 in the UK. Water Research, 200, 117214. https://doi.org/10.1016/j.watres.2021.117214

Juel, M. A. I., Stark, N., Nicolosi, B., Lontai, J., Lambirth, K., Schlueter, J., Gibas, C., & Munir, M. (2021). Performance evaluation of virus concentration methods for implementing SARS-CoV-2 wastewater based epidemiology emphasizing quick data turnaround. Science of the Total Environment, 801, 149656. https://doi.org/10.1016/j.scitotenv.2021.149656

Kumar, M., Joshi, M., Patel, A. K., & Joshi, C. G. (2021). Unravelling the early warning capability of wastewater surveillance for COVID-19: A temporal study on SARS-CoV-2 RNA detection and need for the escalation. Environmental Research, 196(February), 110946. https://doi.org/10.1016/j.envres.2021.110946

Kumar, M., Patel, A. K., Shah, A. V., Raval, J., Rajpara, N., Joshi, M., & Joshi, C. G. (2020). First proof of the capability of wastewater surveillance for COVID-19 in India through detection of genetic material of SARS-CoV-2. Science of the Total Environment, 746, 141326. https://doi.org/10.1016/j.scitotenv.2020.141326

McHugh, M. L. (2012). Lessons in biostatistics interrater reliability : the kappa statistic. Biochemica Medica, 22(3), 276–282. https://hrcak.srce.hr/89395

Medema, G., Heijnen, L., Elsinga, G., Italiaander, R., & Medema, G. (2020). Presence of SARS-Coronavirus-2 in sewage. Methods Sewage samples. MedRxiv. https://doi.org/10.1101/2020.03.29.20045880

Miyani, B., Fonoll, X., Norton, J., Mehrotra, A., & Xagoraraki, I. (2020). SARS-CoV-2 in Detroit Wastewater. Journal of Environmental Engineering, 146(11), 06020004. https://doi.org/10.1061/(asce)ee.1943-7870.0001830

Murakami, M., Hata, A., Honda, R., & Watanabe, T. (2020). Letter to the Editor: Wastewater-Based Epidemiology Can Overcome Representativeness and Stigma Issues Related to COVID-19. Environmental Science and Technology, 54(9), 5311. https://doi.org/10.1021/acs.est.0c02172

Nemudryi, A., Nemudraia, A., Wiegand, T., Vanderwood, K. K., Wilkinson, R., Wiedenheft, B., Nemudryi, A., Nemudraia, A., Wiegand, T., Surya, K., Buyukyoruk, M., & Cicha, C. (2020). Report Temporal Detection and Phylogenetic Assessment of SARS-CoV-2 in Municipal Wastewater ll ll Temporal Detection and Phylogenetic Assessment of SARS-CoV-2 in Municipal Wastewater. Cell Reports Medicine, 1(6), 100098. https://doi.org/10.1016/j.xcrm.2020.100098

Obermeier, P., Muehlhans, S., Hoppe, C., Karsch, K., Tief, F., Seeber, L., Chen, X., Conrad, T., Boettcher, S., Diedrich, S., & Rath, B. (2016). Enabling Precision Medicine With Digital Case Classification at the Point-of-Care. EBioMedicine, 4, 191–196. https://doi.org/10.1016/j.ebiom.2016.01.008

Olesen, S. W., Imakaev, M., & Duvallet, C. (2021). Making waves: Defining the lead time of wastewater-based epidemiology for COVID-19. Water Research, 202, 117433. https://doi.org/10.1016/j.watres.2021.117433

Peccia, J., Zulli, A., Brackney, D. E., Grubaugh, N. D., Kaplan, E. H., Casanovas-massana, A., Ko, A. I., Malik, A. A., Wang, D., Wang, M., Warren, J. L., Weinberger, D. M., Arnold, W., & Omer, S. B. (2020). Measurement of SARS-CoV-2 RNA in wastewater tracks community infection dynamics. Nature Biotechnology, 38(October). https://doi.org/10.1038/s41587-020-0684-z

Pecson, B. M., Darby, E., Haas, C. N., Amha, Y. M., Bartolo, M., Danielson, R., Dearborn, Y., Di Giovanni, G., Ferguson, C., Fevig, S., Gaddis, E., Gray, D., Lukasik, G., Mull, B., Olivas, L., Olivieri, A., Qu, Y., & Sars-Cov-2 Interlaboratory Consortium. (2021). Reproducibility and sensitivity of 36 methods to quantify the SARS-CoV-2 genetic signal in raw wastewater: Findings from an interlaboratory methods evaluation in the U.S. Environmental Science: Water Research and Technology, 7(3), 504–520. https://doi.org/10.1039/d0ew00946f

Randazzo, W., Truchado, P., Cuevas-Ferrando, E., Simón, P., Allende, A., & Sánchez, G. (2020). SARS-CoV-2 RNA in wastewater anticipated COVID-19 occurrence in a low prevalence area. Water Research, 181. https://doi.org/10.1016/j.watres.2020.115942

Saguti, F., Magnil, E., Enache, L., Patzi, M., Johansson, A., Lumley, D., Davidsson, F., Dotevall, L., Mattsson, A., Trybala, E., Lagging, M., Lindh, M., Gisslén, M., Brezicka, T., Nyström, K., & Norder, H. (2021). Surveillance of wastewater revealed peaks of SARS-CoV-2 preceding those of hospitalized patients with COVID-19. 189. https://doi.org/10.1016/j.watres.2020.116620

Sherchan, S. P., Shahin, S., Ward, L. M., Tandukar, S., Aw, T. G., Schmitz, B., Ahmed, W., & Kitajima, M. (2020). Science of the Total Environment First detection of SARS-CoV-2 RNA in wastewater in North America : A study in Louisiana, USA. Science of the Total Environment, 743, 140621. https://doi.org/10.1016/j.scitotenv.2020.140621

Weidhaas, J., Aanderud, Z. T., Roper, D. K., Vanderslice, J., Brown, E., Ostermiller, J., Hoffman, K., Jamal, R., Heck, P., Zhang, Y., Torgersen, K., Vander, J., & Lacross, N. (2021). Science of the Total Environment Correlation of SARS-CoV-2 RNA in wastewater with COVID-19 disease burden in sewersheds. Science of the Total Environment, 775, 145790. https://doi.org/10.1016/j.scitotenv.2021.145790

Westhaus, S., Weber, F., Schiwy, S., Linnemann, V., Brinkmann, M., Widera, M., Greve, C., Janke, A., Hollert, H., Wintgens, T., & Ciesek, S. (2021). Science of the Total Environment Detection of SARS-CoV-2 in raw and treated wastewater in Germany – Suitability for COVID-19 surveillance and potential transmission risks. Science of the Total Environment, 751, 141750. https://doi.org/10.1016/j.scitotenv.2020.141750

WHO. (2020a). Laboratory biosafety guidance related to coronavirus disease (COVID-19). May, 1–11.

WHO. (2020b). Responding to community spread of COVID-19. Interim Guidance 7 March, March, 1–6. https://www.who.int/publications/i/item/responding-to-community-spread-of-covid-19

Wurtzer, S., Marechal, V., Mouchel, J.-M., Maday, Y., Teyssou, R., Richard, E., Almayrac, J. L., & Moulin, L. (2020). Evaluation of lockdown impact on SARS-CoV-2 dynamics through viral genome quantification in Paris wastewaters. MedRxiv, 2020.04.12.20062679. https://doi.org/10.1101/2020.04.12.20062679

Yan Bai, Yao, L., TaoWei Tian, F., Jin, D.-Y., Chen, L., & Meiyun Wang. (2020). Presumed Asymptomatic Carrier Transmission of COVID-19. JAMA, 382(13), 1199–1207. https://doi.org/10.1056/nejmoa2001316

Zhao, L., Atoni, E., Nyaruaba, R., Du, Y., Zhang, H., Donde, O., Huang, D., Xiao, S., Ren, N., Ma, T., Shu, Z., Yuan, Z., Tong, L., & Xia, H. (2021). Environmental surveillance of SARS-CoV-2 RNA in wastewater systems and related environments in Wuhan : April to May of 2020. Journal of Environmental Sciences, 0–17. https://doi.org/10.1016/j.jes.2021.05.005

