## Supplementary Files for "Tracking the temporal variation of COVID-19 surges through wastewater-based epidemiology during the peak of the pandemic: a six-month long study in Charlotte, North Carolina"

* These authors contributed equally

**Supplementary file**

**(a)**

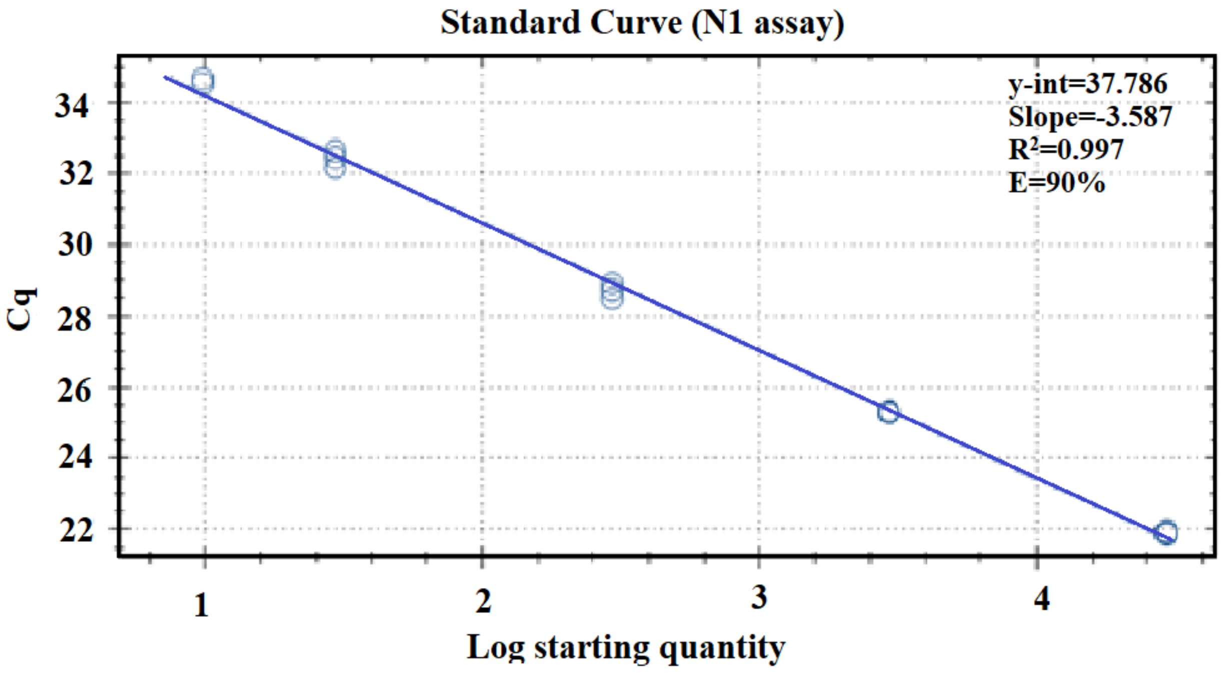

(b)

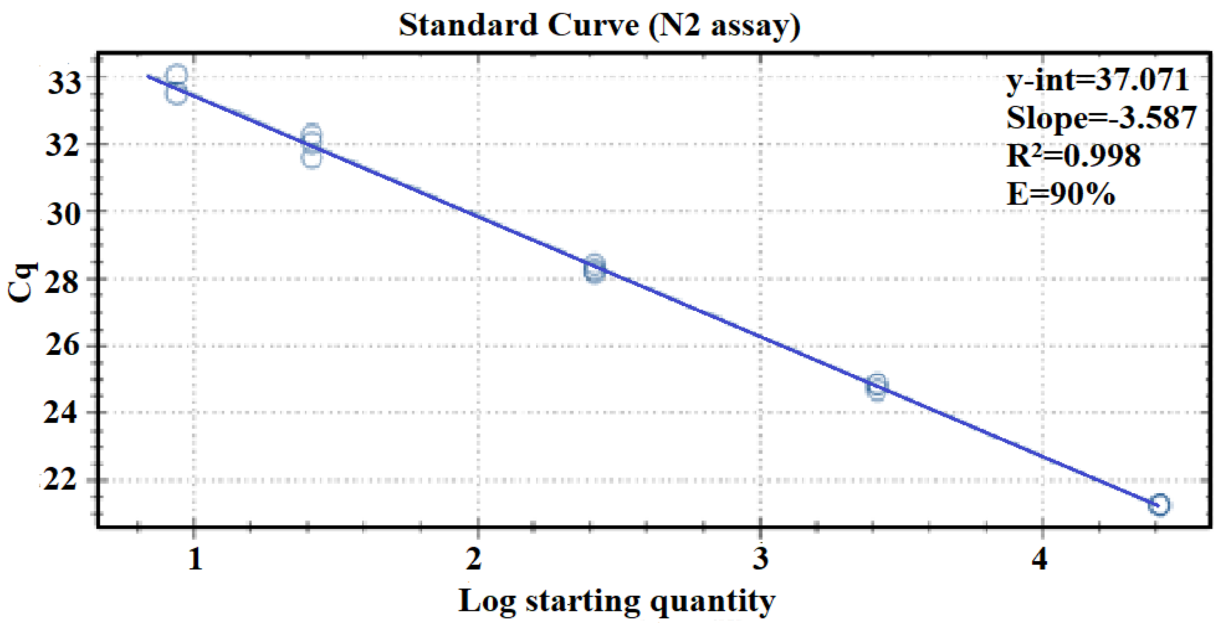


**Fig. S1.** Standard curve for (a) N1 and (b) N2 assay using workflow 1.

**Table S1.** Primers and probe sequences utilized in this WBE study.

| Assay | Primer/Probe | Sequences | Reference |
|---|---|---|---|
| **CDC N1** | Forward | GACCCCAAAATCAGCGAAAT | [1]CDC (2020) |
| | Reverse | TCTGGTTACTGCCAGTTGAATCTG | |
| | Probe | FAM-ACCCCGCATTACGTTTGGTGGACC-BHQ1 | |
| **CDC N2** | Forward | TTACAAACATTGGCCGCAAA | [1]CDC (2020) |
| | Reverse | GCGCGACATTCCGAAGAA | |
| | Probe | FAM-ACAATTTGCCCCCAGCGCTTCAG-BHQ1 | |
| **BCoV** | Forward | CTGGAAGTTGGTGGAGTT | [2]Decaro et al., 2008 |
| | Reverse | ATTATCGGCCTAACATACATC | |
| | Probe | FAM-CCTTCATATCTATACACATCAAGTTGTT-BHQ1 | |

| | | | |
|---|---|---|---|
| **Hep G** | Forward | CGG CCA AAA GGT GGT GGA TG | [3]Schlueter et al., 1996 |
| | Reverse | CGA CGA GCC TGA CGT CGG G | |
| | Probe | HEX-AGG TCC CTC TGG CGC TTG TGG CGA G-BHQ1 | |

**Table S2.** Average recovery efficiency of BCoV and HepG for workflow 1

| | Workflow 1 | |
|---|---|---|
| **Plant** | **BCoV recovery (%)** | **Hep G recovery (%)** |
| **A** | 21.5±15 | 41.31±18.1 |
| **B** | 22±23 | 43.4±21 |
| **C** | 20.2±24 | 38±22.7 |
| **D** | 30.9±29 | 44±17.4 |

**Table S3.** Average recovery efficiency of BCoV and HepG for workflow 2

| | Workflow 2 | |
|---|---|---|
| **Plant** | **BCoV recovery (%)** | **Hep G recovery (%)** |
| **A** | 24.76±20 | 26.22±15.5 |
| **B** | 22.3±17 | 17.3±11.5 |
| **C** | 23.3±18 | 24±13.6 |
| **D** | 26±25 | 29.8±19.6 |

**Table S4.** Inhibition control data for N1 gene for workflow 1 (RT-qPCR)

| Date | Plant | Undiluted | | | | Diluted (1:5) | | | |
|---|---|---|---|---|---|---|---|---|---|
| | | Cq | | | | Cq | | | |
| | | Rep#1 | Rep#2 | Rep#3 | Avg. Cq | Rep#1 | Rep#2 | Rep#3 | Avg. Cq |
| 11/23/2020 | A | 33.67 | 31.89 | 32.34 | 32.63 | 35.54 | ND | 36.29 | 35.9 |
| | B | 34.24 | 33.37 | 33.92 | 33.77 | ND | ND | ND | NA |
| | C | 34.7 | 32.96 | 34.94 | 34.20 | ND | ND | ND | NA |
| 11/25/2020 | A | 33.24 | 33.27 | 33.33 | 33.28 | ND | ND | ND | NA |
| | B | 36.07 | 33.31 | 33.92 | 34.43 | ND | ND | ND | NA |
| | C | 32.00 | 32.54 | 32.01 | 32.18 | 36.85 | ND | ND | NA |
| | D | ND | ND | ND | NA | ND | ND | ND | NA |
| 11/30/2020 | A | 32.9 | 32.92 | 32.19 | 32.67 | 36.00 | ND | ND | NA |
| | B | 31.87 | 33.3 | 33.07 | 32.74 | ND | ND | 35.44 | 35.44 |
| | C | 32.33 | 32.1 | 32.24 | 32.22 | 36.24 | ND | ND | 36.24 |

| Date | Plant | Undiluted | | | | Diluted (1:3) | | | |
|---|---|---|---|---|---|---|---|---|---|
| | | Cq | | | | Cq | | | |
| | | Rep#1 | Rep#2 | Rep#3 | Avg. Cq | Rep#1 | Rep#2 | Rep#3 | Avg. Cq |
| 7/1/2020 | A | 33.93 | 32.92 | - | 33.42 | 33.87 | 35.55 | 35.87 | 35.10 |
| | B | ND | ND | - | NA | ND | ND | ND | NA |
| | C | 34.17 | 35.16 | - | 34.66 | 34.41 | 36.47 | ND | 35.44 |
| | D | 34.61 | 32.52 | - | 33.57 | 33.19 | ND | 34.74 | 33.97 |
| 7/8/2020 | B | 37.81 | 35.45 | - | 36.63 | 37.80 | ND | ND | 37.63 |
| | C | 36.22 | 34.36 | - | 35.29 | ND | 35.97 | ND | 35.97 |

| | | | | | | | | | |
|---|---|---|---|---|---|---|---|---|---|
| | D | ND | ND | - | NA | ND | ND | ND | NA |
| 7/22/2020 | A | 33.07 | 32.10 | - | 32.59 | ND | 33.01 | 34.02 | 34.81 |
| | B | 36.83 | 36.12 | - | 36.47 | ND | ND | ND | NA |
| | C | 37.21 | 32.47 | - | 34.84 | ND | 35.28 | ND | 35.28 |
| | D | 37.05 | 34.71 | - | 35.88 | ND | ND | ND | NA |
| 7/29/2020 | A | ND | ND | - | NA | ND | ND | ND | NA |
| | B | 33.73 | ND | - | NA | ND | ND | ND | NA |
| | C | 35.12 | 33.69 | - | 34.41 | 35.96 | ND | ND | 35.96 |
| | D | ND | ND | - | NA | 35.36 | 35.29 | 33.94 | 34.86 |

**Table S5.** Inhibition control data for N2 gene for workflow 2 (ddPCR)

| Date | Plant | Undiluted | | | | Diluted (1:3) | | | |
|---|---|---|---|---|---|---|---|---|---|
| | | Cq | | | | Cq | | | |
| | | Rep#1 | Rep#2 | Rep#3 | Avg. Cq | Rep#1 | Rep#2 | Rep#3 | Avg. Cq |
| 7/1/2020 | A | 35.23 | 34.22 | 37.43 | 35.63 | 35.29 | 34.76 | ND | 35.03 |
| | B | ND | ND | ND | NA | ND | ND | ND | NA |
| | C | ND | 37.15 | 36.06 | 36 | 34.61 | ND | 37.33 | 35.97 |
| | D | ND | 41.17 | 39.34 | 40.25 | 35.53 | ND | 33.85 | 34.69 |
| 7/8/2020 | B | ND | ND | ND | NA | ND | ND | ND | NA |
| | C | ND | ND | 35.39 | 35.39 | ND | ND | ND | NA |
| | D | ND | ND | ND | NA | ND | ND | ND | NA |
| 7/22/2020 | A | 33.60 | 33.79 | 34.33 | 35.91 | ND | 35.94 | ND | 35.94 |
| | B | ND | ND | ND | NA | ND | ND | ND | NA |
| | C | 33.97 | 36.93 | 34.29 | 35.06 | ND | 35.94 | ND | 35.94 |

| | | | | | | | | | |
|---|---|---|---|---|---|---|---|---|---|
| | D | ND | ND | ND | NA | ND | 36.74 | ND | 36.74 |
| 7/29/2020 | A | 36.75 | ND | ND | 36.75 | ND | ND | ND | NA |
| | B | ND | ND | ND | NA | ND | ND | 39.80 | 39.80 |
| | C | 36.46 | 34.16 | 36.52 | 35.71 | ND | ND | 36.43 | 36.43 |
| | D | ND | ND | ND | NA | 37.93 | ND | ND | NA |

| **Date** | **Plant** | **Undiluted** | | | | **Diluted (1:2)** | | | |
|---|---|---|---|---|---|---|---|---|---|
| | | | **Cq** | | **Avg. Cq** | | **Cq** | | **Avg. Cq** |
| 7/15/2020 | A | ND | ND | ND | NA | ND | ND | ND | NA |
| | C | ND | ND | ND | NA | ND | ND | ND | NA |
| 8/19/2020 | C | ND | ND | ND | NA | ND | ND | ND | NA |
| | D | ND | ND | ND | NA | ND | ND | ND | NA |
| 8/26/2020 | A | ND | ND | ND | NA | ND | ND | ND | NA |
| | B | ND | ND | ND | NA | ND | ND | ND | NA |
| | C | ND | ND | ND | NA | ND | ND | ND | NA |
| 9/23/2020 | A | ND | ND | ND | NA | ND | ND | ND | NA |
| | B | ND | ND | ND | NA | ND | ND | ND | NA |
| | C | ND | ND | ND | NA | ND | ND | ND | NA |
| | D | ND | ND | ND | NA | ND | ND | ND | NA |
| 9/30/2020 | C | ND | ND | ND | NA | ND | ND | ND | NA |
| 10/7/2020 | A | ND | ND | ND | NA | ND | ND | ND | NA |
| | B | ND | ND | ND | NA | ND | ND | ND | NA |
| | C | ND | ND | ND | NA | ND | ND | ND | NA |
| | D | ND | ND | ND | NA | ND | ND | ND | NA |

| | | | | | | | | | |
|---|---|---|---|---|---|---|---|---|---|
| 11/23/2020 | A | ND | ND | ND | NA | 33.22 | 33.65 | ND | 33.44 |
| | B | ND | ND | ND | NA | 35.55 | 34.52 | 34.04 | 34.70 |
| | C | ND | ND | ND | NA | 33.51 | 33.41 | ND | 33.46 |
| 11/25/2020 | A | ND | ND | ND | NA | 34.26 | 33.78 | 33.9 | 33.98 |
| | B | ND | ND | ND | NA | ND | 35.16 | 35.67 | 35.42 |
| | C | ND | ND | ND | NA | 33.69 | 34.13 | 33.38 | 33.73 |
| | D | ND | ND | ND | NA | ND | ND | ND | NA |
| 11/30/2020 | A | ND | ND | ND | NA | ND | 33.66 | 33.5 | 33.58 |
| | B | ND | ND | ND | NA | 33.78 | 33.74 | ND | 33.76 |
| | C | ND | ND | ND | NA | ND | 33.16 | 33.97 | 33.57 |
| 12/2/2020 | A | ND | ND | ND | NA | 31.37 | 31.21 | 32.03 | 31.54 |
| | B | ND | ND | ND | NA | 32.93 | 33.51 | 33.01 | 33.15 |
| | C | ND | ND | ND | NA | 33.64 | 33.1 | ND | 33.37 |
| | D | ND | ND | ND | NA | ND | ND | ND | NA |
| 12/7/2020 | A | ND | ND | ND | NA | 32.69 | 33.15 | 32.96 | 32.93 |
| | B | ND | ND | ND | NA | 34.18 | 33.82 | ND | 34 |
| | C | ND | ND | ND | NA | ND | 33.18 | 35.03 | 34.12 |

- Not detected; NA- Not applicable

**Table S6.** Inhibiton control data for workflow 2 (ddPCR)

| | Inhibition efficiency (%) | | | |
|---|---|---|---|---|
| **Date** | **Plant A** | **Plant B** | **Plant C** | **Plant D** |
| 6/24/2020 | 101 | 101 | 100 | - |
| 7/1/2020 | 101 | 101 | 104 | 99 |
| 7/8/2020 | 96 | 104 | 98 | 101 |
| 7/15/2020 | 102 | 98 | 100 | 97 |
| 7/22/2020 | 99 | 98 | 103 | 99 |
| 7/29/2020 | 98 | 99 | 97 | 98 |
| 8/5/2020 | 94 | 98 | NT | 98 |
| 8/12/2020 | 100 | 98 | 100 | 97 |
| 8/19/2020 | 101 | 98 | 97 | 102 |
| 8/26/2020 | 97 | 100 | 99 | 97 |
| 9/2/2020 | 97 | 96 | 99 | 94 |
| 9/9/2020 | 95 | 92 | - | 98 |
| 9/15/2020 | - | 98 | 89 | - |
| 9/30/2020 | 98 | 97 | 94 | 91 |
| 10/7/2020 | 96 | 90 | 94 | 95 |
| 10/14/2020 | 98 | 95 | 94 | 97 |

| | | | | |
|---|---|---|---|---|
| 10/28/2020 | 96 | 95 | 101 | 93 |
| 11/2/2020 | 98 | 105 | 100 | - |
| 11/4/2020 | 96 | 93 | - | - |
| 11/11/2020 | 93 | 94 | 101 | 96 |
| 11/16/2020 | 93 | 100 | 94 | - |
| 11/18/2020 | 95 | 93 | 97 | - |
| 11/23/2020 | 97 | 97 | 96 | - |
| 11/25/2020 | 94 | 93 | 94 | 96 |
| 11/30/2020 | 96 | 101 | 97 | - |
| 12/7/2020 | 101 | 96 | 94 | - |
| 12/9/2020 | 90 | 95 | 95 | 91 |
| 12/14/2020 | - | 95 | 91 | - |

**Table S7a.** Detailed reagents and methods utilized in workflow 2

**Kingfisher Magnetic bead extraction**

| Quantity | Description | Vendor | Catalog Number |
|---|---|---|---|
| 1 | Tip comb | Fisher Sci. | 22-387-029 |
| 7 | 96 well, DWP | Fisher Sci. | 22-387-031 |
| 800 µL | Wash Buffer 1 | Biomerieux | 280130 |
| 1000 µL | Wash Buffer 2 | Biomerieux | 280131 |
| 500 µL | Wash Buffer 3 | Biomerieux | 280132 |
| 50 µL | Magnetic silica | Biomerieux | 280134 |
| 100 µL | Buffer AE | Life Technologies | 19077 |

**Extraction protocol for workflow 2**

***Lysate Plate***

- Dispense up to 950 µL of supernatant of lysed sample into a 96 Deep Well plate containing 50 mL of magnetic beads.
- Incubate 96 well Deep Well plate containing sample and Magnetic Beads for 10 minutes at room temperature.
- While incubating, set up other plates.

***Wash 1 Plate***

- Dispense 400 µL of Wash Buffer 1 into each well

***Wash 2 Plate***

- Dispense 400 µL of Wash Buffer 1 into each well

***Wash 3 Plate***

- Dispense 500 µL of Wash Buffer 2 into each well

***Wash 4 Plate***

- Dispense 500 µL of Wash Buffer 2 into each well

***Wash 5 Plate***

- Dispense 500 µL of Wash Buffer 3 into each well

***Eluate Plate***

- Dispense 100 µL of AE Elution Buffer into each well

***Tip Comb Plate***

- Place 96 DW tip comb into plate

After 10-minutes of incubation, plates were added to the Kingfisher Instrument as the program directed.

**Table S7b.** Sample preparation for Reverse Transcription

| Quantity | Description | Vendor | Catalog Number |
|---|---|---|---|
| 50 µL | Superscript VILO IV MM | Fisher Sci. | 11756050 |
| 1 | Foil seal | BioRad | 1814040 |
| 1 | 96 well PCR plate | BioRad | 12001925 |
| 1µL | Mouse Lung RNA | BioChain | R1334152-50 |

| RT MasterMIX | µL |
|---|---|
| Volume per rxn | 70 |
| Superscript IV VILO 5X | 14 |
| RNA template | 35 |
| Mouse Lung RNA | 1 |
| Water | 20 |

### References


(1) CDC, (2020). **(https://www.cdc.gov/healthywater/surveillance/wastewater-surveillance/testing-methods.html)**

(2) Decaro, N.; Elia, G.; Campolo, M.; Desario, C.; Mari, V.; Radogna, A.; Colaianni, M. L.; Cirone, F.; Tempesta, M.; Buonavoglia, C. Detection of Bovine Coronavirus Using a TaqMan-Based Real-Time RT-PCR Assay. *J. Virol. Methods* 2008, *151* (2), 167–171. https://doi.org/10.1016/j.jviromet.2008.05.016.

(3) Schlueter, V.; Schmolke, S.; Stark, K.; Hess, G.; Ofenloch-Haehnle, B.; Engel, A. M. Reverse Transcription-PCR Detection of Hepatitis G Virus. *J. Clin. Microbiol.* 1996, *34* (11), 2660–2664. https://doi.org/10.1128/jcm.34.11.2660-2664.1996.
